# Assessing yellow fever outbreak potential and implications for vaccine strategy

**DOI:** 10.1101/2023.12.19.23300139

**Authors:** Keith Fraser, Arran Hamlet, Kévin Jean, Daniel Garkauskas Ramos, Alessandro Romano, Jennifer Horton, Laurence Cibrelus, Neil Ferguson, Katy A.M. Gaythorpe

## Abstract

**Background:** Yellow fever (YF), a vector-borne viral hemorrhagic fever, is endemic in tropical regions of Africa and South America, with large vaccination programmes being used for control. However, significant outbreaks have occurred in recent years. Data on infection rates and seroprevalence is often sparse, requiring robust mathematical models to estimate the burden of yellow fever. In particular, modelling is required to estimate the risk of outbreaks and inform policy decisions regarding the targeting of vaccination.

**Methods:** We present a dynamic, stochastic model of YF transmission which uses environmental covariates to estimate the force of infection due to spillover from the sylvatic (non-human primate) reservoir and the basic reproduction number for human-to-human transmission. We examine the potential for targets identified by the World Health Organization EYE Strategy (50%, 60% or 80% vaccination coverage in 1-60 year olds) to achieve different threshold values for the effective reproduction number. Threshold values are chosen to reflect the potential for seasonal and/or climatic variation in YF transmission even in a scenario where vaccination lowers the median reproduction number below 1.

**Results:** Based on parameter estimates derived from epidemiological data, it is found that the 2022 EYE Strategy target coverage is sufficient to reduce the static averaged annual effective reproduction number *R* below 1 across most or all regions in Africa depending on the effectiveness of reported vaccinations, but insufficient to reduce it below 0.5 and thereby eliminate outbreaks in areas with high seasonal range. Coverage levels aligned with the 2026 targets are found to significantly decrease the proportion of regions where *R* is greater than 0.5.

## 1 Introduction

Yellow fever (YF), a viral hemorrhagic fever spread by insect vectors, has a serious impact in tropical regions in Africa and South America ^1^. The estimated median fatality rate is 39% ^2^ in cases showing severe symptoms (median estimate 12% of all cases ^3^). Due to the existence of a sylvatic reservoir in non-human primates (NHPs), full eradication is not possible, and vaccination is the main form of control.

Eradicating large, self-sustaining outbreaks with significant human-to-human transmission is a key YF policy goal. In recent years, there have been a number of YF outbreaks ^4^. The projection and prevention of outbreaks is particularly important in scenarios where YF may be introduced into a region with no previous history of YF burden, putting un-vaccinated, immunologically näıve populations at risk ^5^. Such introduction of YF into new regions is expected to become more probable due to the effects of climate change ^6^. Previous work on numerical modelling of YF transmission ^7–10^ has generally focused on overall burden over long periods of time rather than on the frequency and size of such outbreaks.

Here, we report on a new transmission model of YF (building on previous work ^7–10)^, informed by available burden data, and calculating parameters via a range of environmental covariates such as temperature suitability for mosquitoes. This model combines transmission of YF from NHPs to humans (via sylvatic mosquito vectors which feed on both) with transmission between humans via human-feeding mosquito vectors rather than defining separate sylvatic and urban (and/or intermediate/savannah) transmission cycles ^11^.

We apply the model to assess the effectiveness of vaccine coverage targets from the World Health Organization Eliminate Yellow Fever Epidemics (EYE) Strategy ^5^ in preventing outbreaks as evidenced by values of the effective reproduction number estimated via Markov chain Monte Carlo sampling using cross-sectional serological survey data and annual case notification data.

## 2 Methods

### 2.1 Model

#### 2.1.1 Transmission modelling

We developed an age-stratified SEIRV model, illustrated in Figure 1. Sylvatic spillover was governed by a static force of infection *λ_S_*, while human-to-human transmission was governed by a basic reproduction number *R*_0_. The force of infection *λ_S_* also captured introduction of infections from external regions. We combined these infection terms to calculate a dynamic force of infection dependent on spillover and human-human transmission and thus project burden.

**Figure 1:**
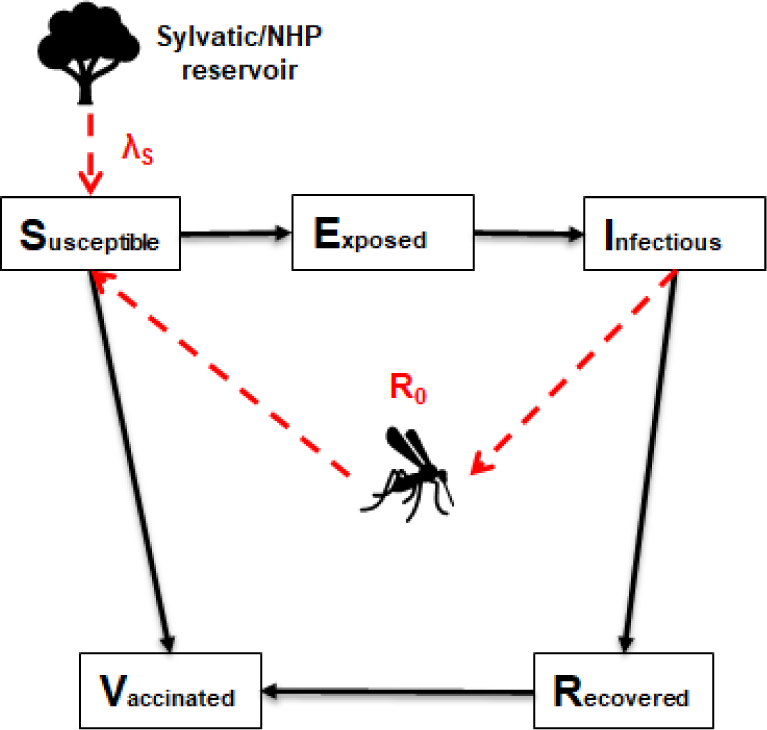
Diagram of SEIRV model of yellow fever transmission in humans showing zoonotic transfer from the sylvatic non-human primate reservoir (mediated by the *λ_S_* parameter) and transmission from infectious humans (mediated by the *R*_0_ parameter)

This model captures both the steady accumulation of YF cases due to sylvatic spillover and the occurrence of outbreaks driven by mosquito-mediated human-to-human transmission. This allows improved differentiation between regions where YF is driven by continuous sylvatic spillover and large outbreaks do not occur, and regions where YF outbreaks are rare due to infrequent sylvatic spillover or introduction from outside the region, but where outbreaks can rapidly grow once nucleated by a spillover event.

Table 1 summarises relevant parameters; the lower section of the table shows the parameters which were estimated using a Markov chain Monte Carlo approach (see section 2.1.2).

**Table 1:**
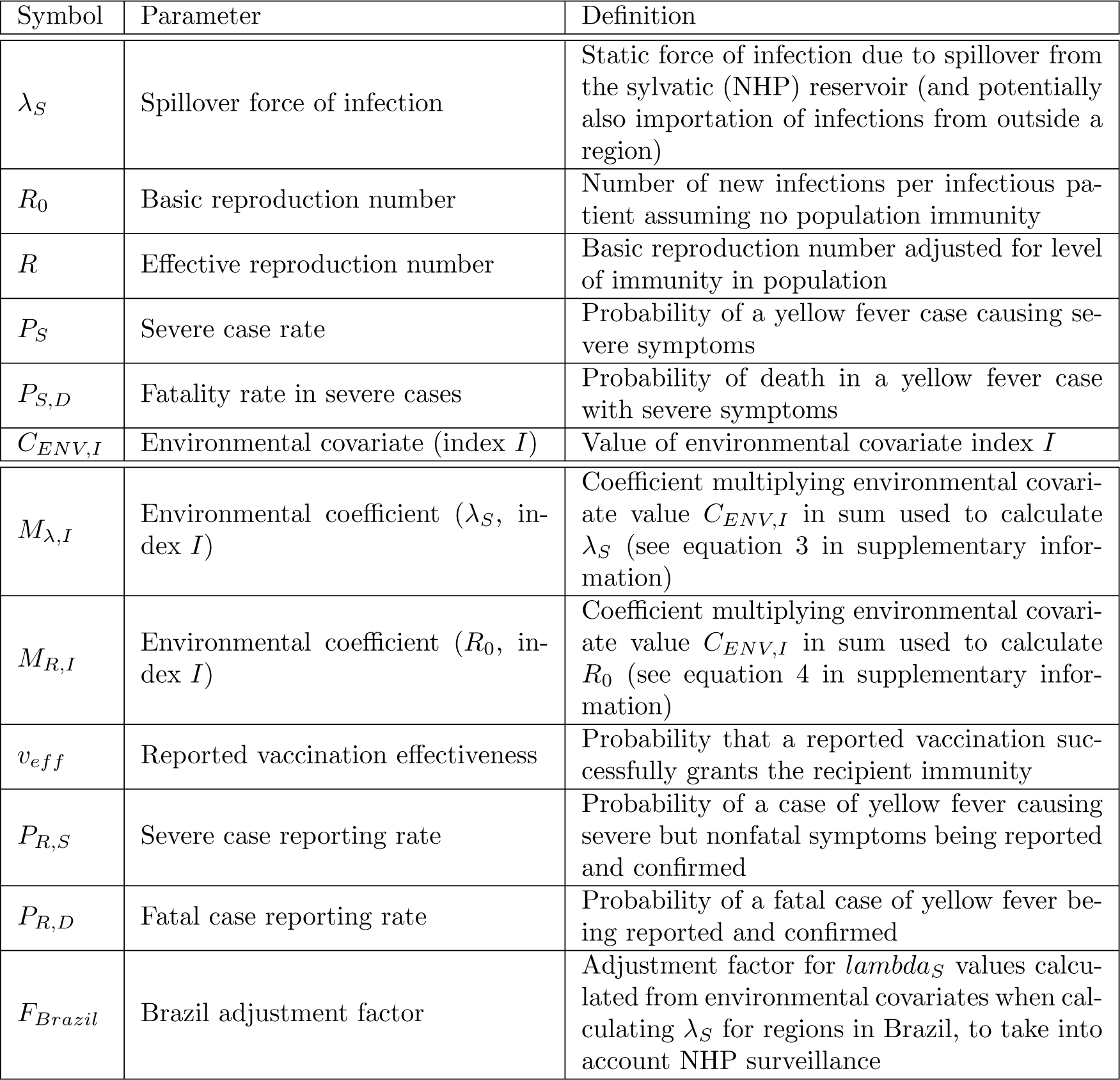
Table of parameters. Lower section shows estimated parameters (see section 2.1.2).

The values of *λ_S_*and *R*_0_ were calculated for a given region from environmental covariates (see section 2.2.2) using a set of coefficients for each (*M_λ,I_* and *M_R,I_*, where *I* is the covariate number) representing the relative importance of each covariate to the value of the parameter. The reported vaccination effectiveness *v_eff_* was incorporated as a variable parameter which encompasses both vaccine efficacy (the probability that an administered vaccination confers immunity on the recipient) and vaccination reporting accuracy (the proportion of reported or estimated vaccinations which were administered).

When generating simulated data on reported cases, additional parameters *P_R,S_*and *P_R,D_*were used, representing the probability of the reporting and confirmation via laboratory testing of infections with severe but non-fatal symptoms, and fatal infections respectively. These two parameters, like the environmental coefficients and reported vaccination effectiveness, were assumed to be constant over all regions and time periods modelled, which may not be the case for real data. As discussed below, the data on reported severe and fatal cases used for parameter estimation was for 6 South American countries over the period 1990-2015. Further case notification data for 11 countries in Africa was used for validation of the estimated model.

An additional parameter *F_Brazil_* was used where relevant to multiply *λ_S_* for regions in the country of Brazil, to take into account surveillance of non-human primates ^12;13^ which is assumed to reduce sylvatic spillover.

For more details of the model, see section 2 of the supplementary information.

#### 2.1.2 Parameter estimation

To estimate the values of the coefficients *M_λ,I_* and *M_R,I_*, along with (where relevant) values of *v_eff_*, *P_R,S_*, *P_R,D_* and *F_Brazil_*, the model was estimated within a Bayesian framework using adaptive Markov Chain Monte Carlo (MCMC) sampling. Multiple chains (4 in the case of the estimation described in this paper) were run for comparative purposes, with samples taken post burn-in given convergence of the chains to provide the posterior predictive distributions. Chain convergence was assessed using the Gelman-Rubin convergence parameter ^14^ in addition to visual inspection. For more details of the estimation method and resulting parameter values, see section 3 of the supplementary information. Distributions of *λ_S_* and *R*_0_ values were calculated from the distributions of estimated values of *M_λ,I_* and *M_R,I_* and the environmental covariates. 1000 sets of parameter values selected at regular intervals from the combined posterior distributions of the 4 Markov chains were also used to compute 1000 sets of serological and annual case data to compare with the observed data. See section 3.1 for the results.

To test the ability of this estimation method to extrapolate epidemiological parameters from sparse data, a set of serological survey and annual case data was simulated. Selected portions of this data were then used to recover the original parameters using the estimation process described above. The results matched the original input parameters closely, with the input values lying within the 95% credible interval of the posterior distribution in all cases. See section 4 in the supplementary information for more details.

### 2.2 Input data

Serological data for parameter estimation was taken from 17 cross-sectional studies carried out across 13 African countries between 1985 and 2019. Annual reported case data for parameter estimation was gathered in 6 South American countries from 1990-2015. Further case data was compiled for 11 countries in Africa to validate the model projections. See section 5.1 in the supplementary information for further details of serological and annual reported case data used for parameter estimation.

#### 2.2.1 Population and vaccination data

Population data was derived from the 2019 United Nations World Population Prospects database ^15^ which was disaggregated sub-nationally using information from Landscan ^16;17^. Vaccination data was derived from historic data on mass-vaccination activities, reactive campaigns, recent preventive mass vaccination campaigns, and routine infant vaccination ^18^ as described in previous work ^10;19^.

#### 2.2.2 Environmental covariate data

The environmental covariates used to calculate *λ_S_* and *R*_0_ from coefficients were as follows ^10^:

- *NHP_combined_*, the richness of NHP species within a region, combining values used ^20^ for the families *cercopithecidae*, *cebidae* and *aotidae*.
- *P_log_*, the natural logarithm of the human population of a region informed by UNWPP and Landscan ^15–17^
- *LC*10, the proportion of a region’s land area covered by grasslands, obtained from MODIS ^21^; this is expected to affect mosquito abundance
- *M_aegypti_*, a covariate with value 1 or 0 representing the presence or absence respectively of *aedes aegypti* mosquitoes, a key vector for urban transmission of yellow fever ^22^; this is used as a proxy for the presence and density of YF-carrying mosquitoes, since other species are not well documented
- *MIR_max_*, maximum middle infrared reflectance ^23^; another covariate reflecting land use affecting mosquito presence and density
- *T_suit,mean_*, suitability of a region’s temperature range for mosquitoes ^8^

In previously reported work ^10^, a covariate selection process was used to select these parameters from a larger list via fitting a generalised linear model to yellow fever occurrence data. In brief, the univariate correlations were calculated between all covariates and YF occurrence, this was used to eliminate parameters with unclear relationship with YF. Then, the remaining covariates were clustered, and the most correlated covariate was chosen from each cluster to contribute step-wise model selection to optimse the Bayesian Information Criterion.

In the case of two low-population island regions (San Andŕes y Providencia in Colombia, An-nobón in Equatorial Guinea), mean temperature data was not available to calculate the temperature suitability index. These regions were therefore not simulated where relevant. Due to the small populations of these regions (under 0.5% of the total populations of the respective countries), their removal was not expected to significantly perturb overall results.

### 2.3 Evaluation of outbreak potential

We used the model to examine the ability of target vaccination coverage levels recommended in the EYE Strategy ^5^ to curtail large yellow fever outbreaks, based on the results above. The relevant quotations are:

> Vaccine coverages greater than 80%, with a 60-80% security threshold, are necessary to interrupt local transmission (human-mosquito-human) of YF virus within a community and to ensure that sporadic unvaccinated cases do not generate additional cases. (EYE Strategy ^5^, page 20)

> By end of 2022: At least 50% of the target population of high-risk countries of Africa has been protected through national preventive mass vaccination campaigns (EYE Strategy^5^, page 41)

The age range of 1-60 was chosen to represent the majority of adults. 50% coverage represents the interim 2022 target (set in 2018) as a minimum or ”worst-case” scenario, with 60% and 80% as the lower and upper bounds of the 2026 target. The criterion used was therefore:

> Are the target coverage levels proposed by the EYE Strategy (50%, 60% or 80% of individuals aged 1-60 vaccinated in African countries with a significant yellow fever burden) sufficient to reduce the effective reproduction number *R* in all of those countries below 1 (thus preventing large-scale outbreaks of human-to-human transmission with exponential growth in infections)?

The basic reproduction number, *R*_0_, was converted to the effective reproduction number, *R*, using the formula *R* = *R*_0_(1-*F_V_* ), with *F_V_*being the successfully vaccinated fraction of the population over all age groups. Note that this will not capture age-based heterogeneity in immunity, as we apply vaccination coverage in the final scenarios evenly across all target age groups.. This allowed the probability that *R ≥* 1 to be calculated for each region at a given vaccination coverage, by calculating the probability that *R*_0_ *≥* 1/(1-*F_V_* ) from the distribution of *R*_0_ values obtained from the results of MCMC parameter estimation. *R*_0_ values were converted into values of the probability that *R*_0_ *≥* 1 for all of the displayed regions, by dividing the number of instances where *R*_0_ *≥* 1 in the MCMC chains post-burn-in by the total number of values.

A more stringent criterion tested was whether the EYE Strategy target coverage is sufficient to reduce *R* to either 0.7 or below or 0.5 or below. The static *R*_0_ used in our model did not take seasonality into account; in a scenario where the static *R*_0_ represents an annual average of a seasonally varying value, the maximum value may (depending on the type of seasonal variation) be significantly higher than this average, hence the more stringent criteria. The work of Codeço et al ^24^ provides an example of large seasonal variations (from below 0.5 to above 2.0) in effective reproduction number for dengue fever, another mosquito-borne flavivirus. In addition to seasonal variation, climate change may result in significant increases in values of some of the environmental covariates used to calculate *R*_0_ used in some regions, such as increased temperature suitability due to changes in median temperature ^8^.

## 3 Results

### 3.1 Parameter estimation based on reported yellow fever data

The combination of African serological survey datasets and South American annual case numbers was used to estimate the values of environmental covariate coefficients used to calculate *λ_S_* and *R*_0_ along with the additional parameters *v_eff_*, *P_R,S_*, *P_R,D_* and *F_Brazil_*. See the supplementary information (section 5) for more details including a plot of likelihood over time displaying the convergence of the 4 Markov chains used (Figure 5), and graphs of obtained parameter value distributions (Figure 6); Table 1 displays the median and 95% CI values of each parameter.

The values of reported vaccination effectiveness *v_eff_* obtained from the estimation process are noteworthy. The median value of *v_eff_*was 60.8% with 95% CI 55.9-65.1%. This is lower than typically estimated values for vaccine efficacy (median 97.5% ^25^), suggesting that there is significant over-estimation in the vaccination coverage data used. This aligns with work examining the sensitivity and specificity of YF vaccination coverage reporting ^26^.

Figures 2 and 3 show the distribution of simulated data based on model output compared with the observed data. For the majority of serological surveys, the modelled results generally overlapped with the confidence intervals for the real values used for estimation (Figure 2). The exceptions were generally from older (pre-2010) surveys and/or for data taken from older patients where the uncertainty in seroprevalence values is higher due to smaller sample sizes.

**Figure 2:**
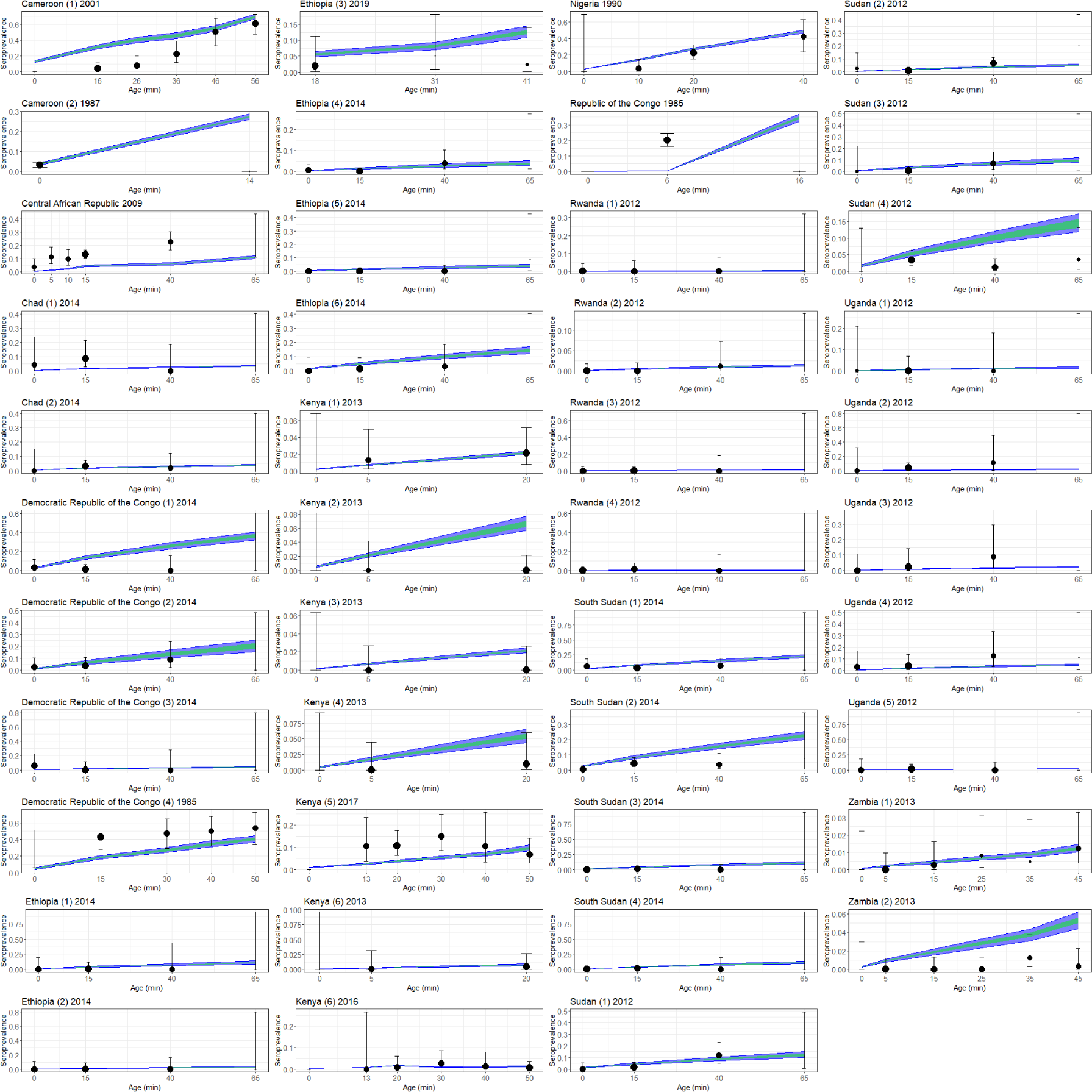
Modelled seroprevalence estimates (coloured regions) compared with real data (black). Model estimates represent a distribution generated from 1000 parameter sets drawn from the postburn-in distribution obtained from 4 Markov chains; green regions show distribution of 50% of results, blue regions show distribution of 95% of results. Error bars on real data obtained from binomial confidence interval calculations using numbers of tested individuals and positive tests. Point sizes represent numbers of tested individuals; x-axis values represent minimum age in age groups.

**Figure 3:**
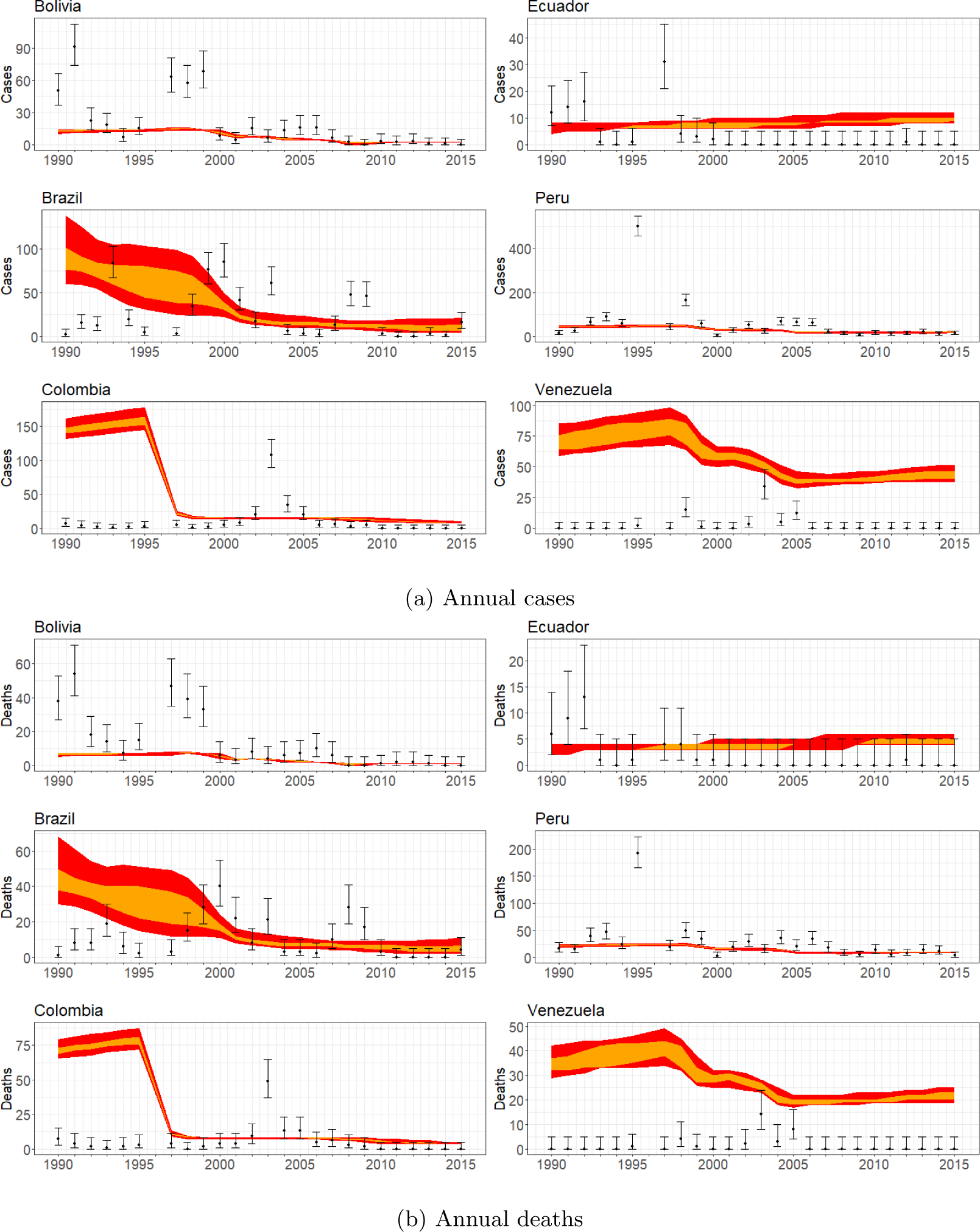
Graphs showing reproduction of selected South American annual case (a) and death (b) data using 1000 parameter sets drawn from post-burn-in distribution obtained from 4 Markov chains. Points show real data (error bars obtained from binomial confidence interval calculations using the population); orange regions show distribution of 50% of results, red regions show distribution of 95% of results.

In the case of the annual reported case and death data from South America (Figure 3), the modelled estimates overlapped with the confidence intervals of the majority of observed values for 4 out of 6 countries (Bolivia, Brazil, Ecuador, Peru). The observed discrepancies are in part due to the unpredictability of large outbreaks in individual years, which these localised spikes in case numbers represent. In the cases of the two countries where the poorest agreement between simulated and observed data is found (Colombia and Venezuela), the discrepancies may be due in part to the very low reported burden (with the majority of years having zero reported cases). Inconsistency in case reporting rates and/or in vaccination reporting may also be responsible - e.g. in the case of Colombia, the lack of the relatively high number of cases projected for 1990-5 may be due to low reporting rates prior to the major vaccination campaign responsible for the drop in projected case rates and/or the effects of increased vaccination appearing earlier than projected. Additional simulated data compared with reported case data from 11 African countries (not used for estimation) is shown in section 5.3 in the supplementary information (Figure 7).

The expected total annual burden of yellow fever across all endemic regions of the world was also calculated using a distribution of 1000 values drawn from the combined posterior distributions of the 4 chains. These calculations covered a total of 734 regions at the first sub-national level (when regions lacking temperature data were excluded) in 45 countries. For comparison with other results in the literature, distributions of values of severe case rate *P_S_*and severe case fatality rate *P_S,D_*were used ^3^ (note that in the MCMC estimation, fixed values of 12% ^3^ and 39% ^2^ were used for these parameters). See the supplementary information (section 5.2) for more details.

The total calculated worldwide deaths in 2018 had a median value of 58,900 with a 95% CrI of 15,100-137,500. The median value was split between 52,100 deaths in Africa and 6,700 in South America. This compared with an estimate of 51,000 worldwide deaths (CrI 31,000-82,000) for the same year in previous work ^10^ and an estimate of 78,000 (CrI 19,000-180,000) for 2013 from earlier work ^7^. Related estimates in other work include a reported estimate of 30,000 annual deaths by the World Health Organization ^27^, and an estimate of 35,000 for 2016 based on other modelling results in the literature ^28^.

Figures 4 and 5 show how the values of *λ_S_* and *R*_0_ derived from the estimated environmental coefficients varied across relevant regions of Africa and South America. The values shown are based on the distribution of values of global burden discussed above; they represent the parameter set which gave the median global burden (when the fixed values of *P_S_* and *P_S,D_* were used) out of the sets tested. The geographical pattern is similar to the maps of total force of infection obtained in previous work ^10^ - the highest values of *λ_S_* were obtained in high-burden regions such as west Africa and the Amazon basin. Regions with negligibly low *λ_S_* generally corresponded to regions with no recorded yellow fever cases since 1984^8^. Values of median *R*_0_ are similarly high in high-burden regions such as west and central Africa and the Amazonas region of Brazil, exceeding 1.0 in many of these areas. These values are significantly lower than obtained from analysis of some recorded outbreaks (e.g. a median of 4.21 and an interquartile range of 2.19 in a review of multiple such estimates ^29^), but as noted in section 2.3, this value represents a year-round average, and seasonal variation and other effects may potentially raise it to much higher values at particular times and/or in particular regions to produce outbreaks with very rapid human-to-human transmission.

**Figure 4:**
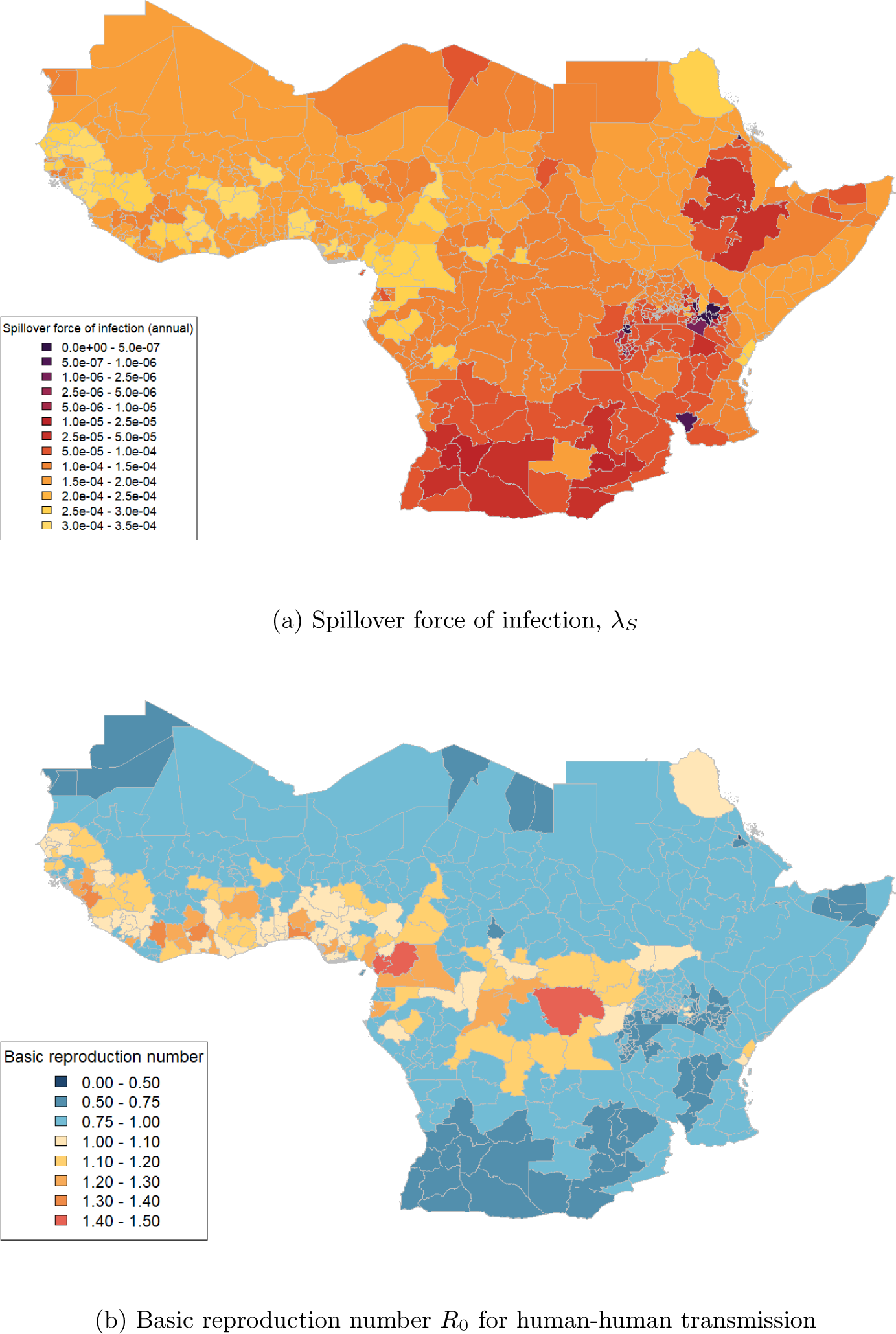
Maps of median projected *λ_S_* (a) and *R*_0_ (b) across 1st-level sub-national administrative regions in selected African countries.

**Figure 5:**
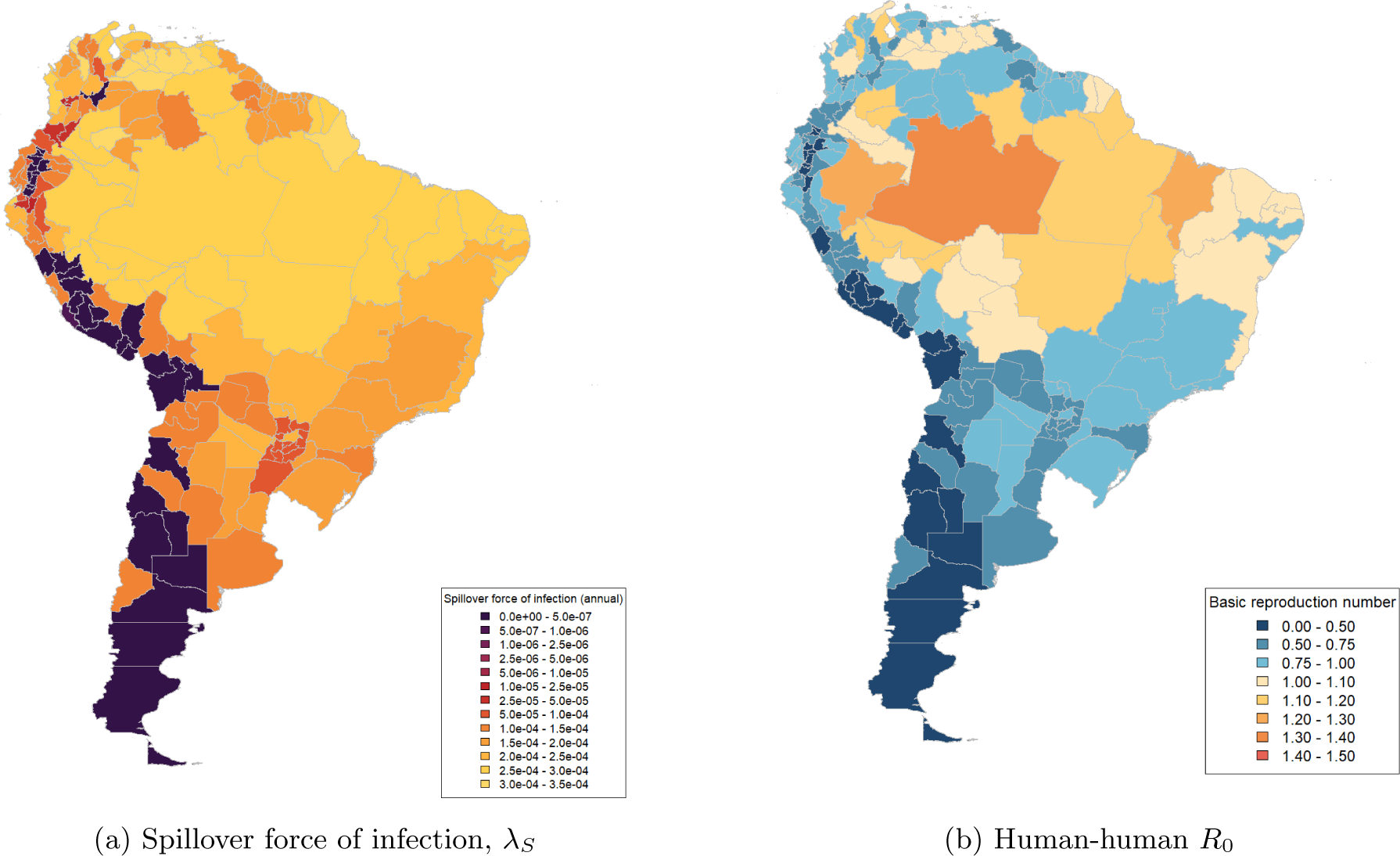
Maps of median projected *λ_S_* (a) and *R*_0_ (b) across 1st-level sub-national administrative regions in selected South American countries. Note that *λ_S_* values shown for Brazil were not adjusted using the parameter *F_B_razil*.

### 3.2 Evaluating vaccination targets

The *R*_0_ values for relevant regions in Africa summarised (as median values) in Figure 4b were converted into values of the probability that *R*_0_ exceeds certain values. The probability that *R*_0_ *≥* 1 was found to be 100% or close to 100% across large sections of west and central Africa and above 5% for many surrounding regions (6a). If the vaccination coverage in individuals aged 1-60 was 50%, in line with the interim 2022 EYE Strategy target (with age groups outside this range assumed to have no vaccination coverage, and no infection-derived immunity assumed), the probability that the effective reproduction number *R ≥* 1 was found to fall to zero for all the regions shown in Figure 6a if all vaccinations were effective (i.e. *v_eff_*= 100%). Therefore, using this criterion, the 2022 EYE Strategy target coverage appears to be sufficient to prevent large-scale outbreaks, assuming that the environmental covariate values used to calculate *R*_0_ (see section 2.2.2) remain unchanged.

**Figure 6:**
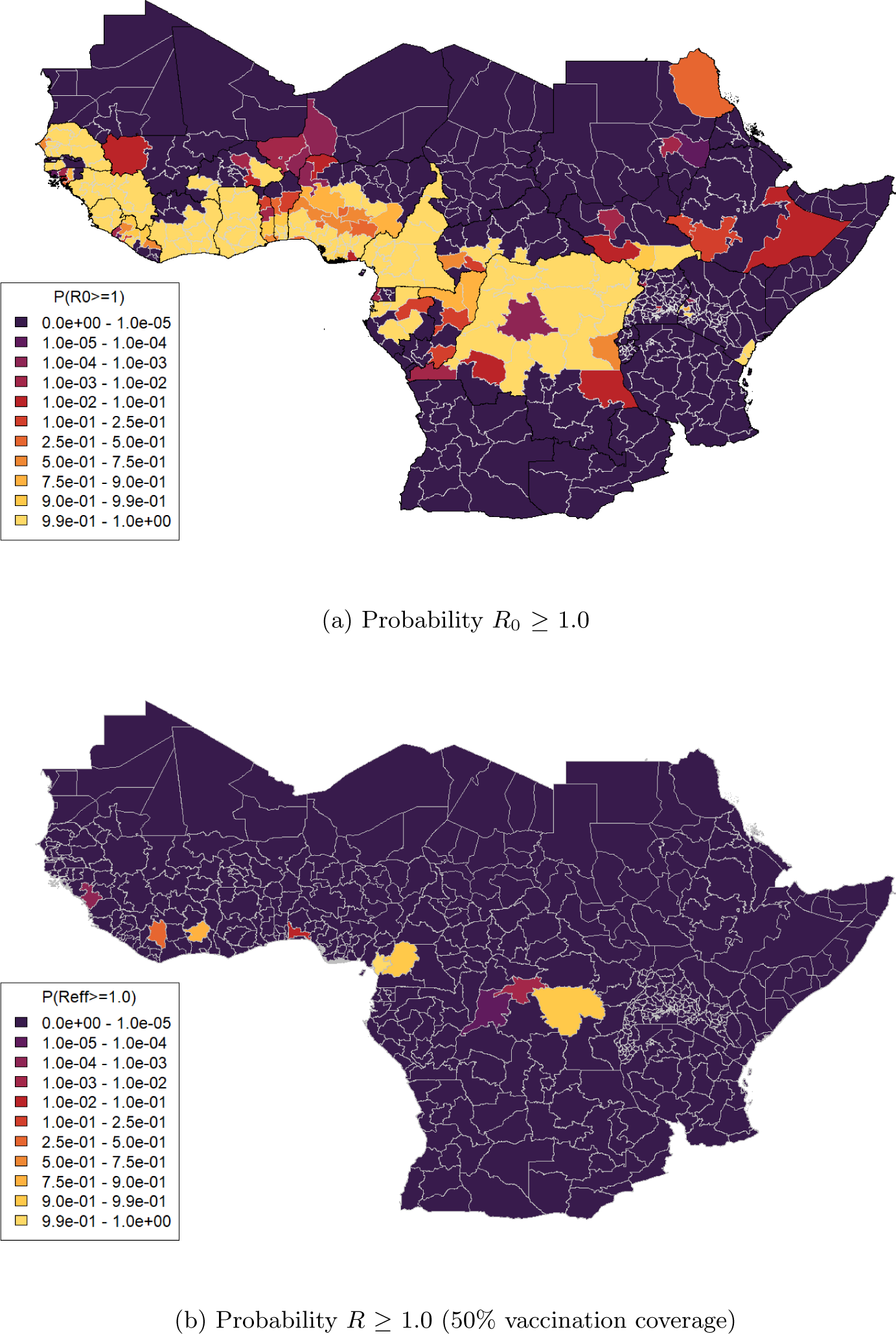
Maps of selected 1st-level sub-national administrative regions in Africa displaying (a) probability that projected *R*_0_ *≥* 1.0 based on data used in Figure 4b b) probability that projected *R ≥* 1.0 for vaccination coverage 50% in 1-60 year olds, based on distributions of values of *R*_0_ and *v_eff_* obtained from MCMC results

Figure 6b shows the probability that *R ≥* 1.0 for the target 50% coverage based on the distribution of *v_eff_* values obtained alongside the *R*_0_ values. With reported vaccination effectiveness taken into account, a small number of regions were found to have non-zero values of the probability that *R ≥* 1.0 and therefore to be at risk of large-scale outbreaks. If the vaccination coverage was raised from 50% to 60% or 80%, representing the 2026 EYE Strategy target range, the number of regions where the probability that *R ≥* 1.0 was over 5% fell to below 5 (for 60%) or zero (for 80%).

For the stricter criterion *R ≥* 0.5 (see section 2.3), many more regions were found to be at risk of large-scale outbreaks. Figures 7a-b show the probability that *R ≥* 0.5 for vaccination coverage values of 50% and 80% respectively. For the lower vaccination coverage, the criterion is not met across the majority of the relevant regions; for the higher value, the proportion of regions where the probability that *R ≥* 0.5 is over 5% goes down, but remains high.

**Figure 7:**
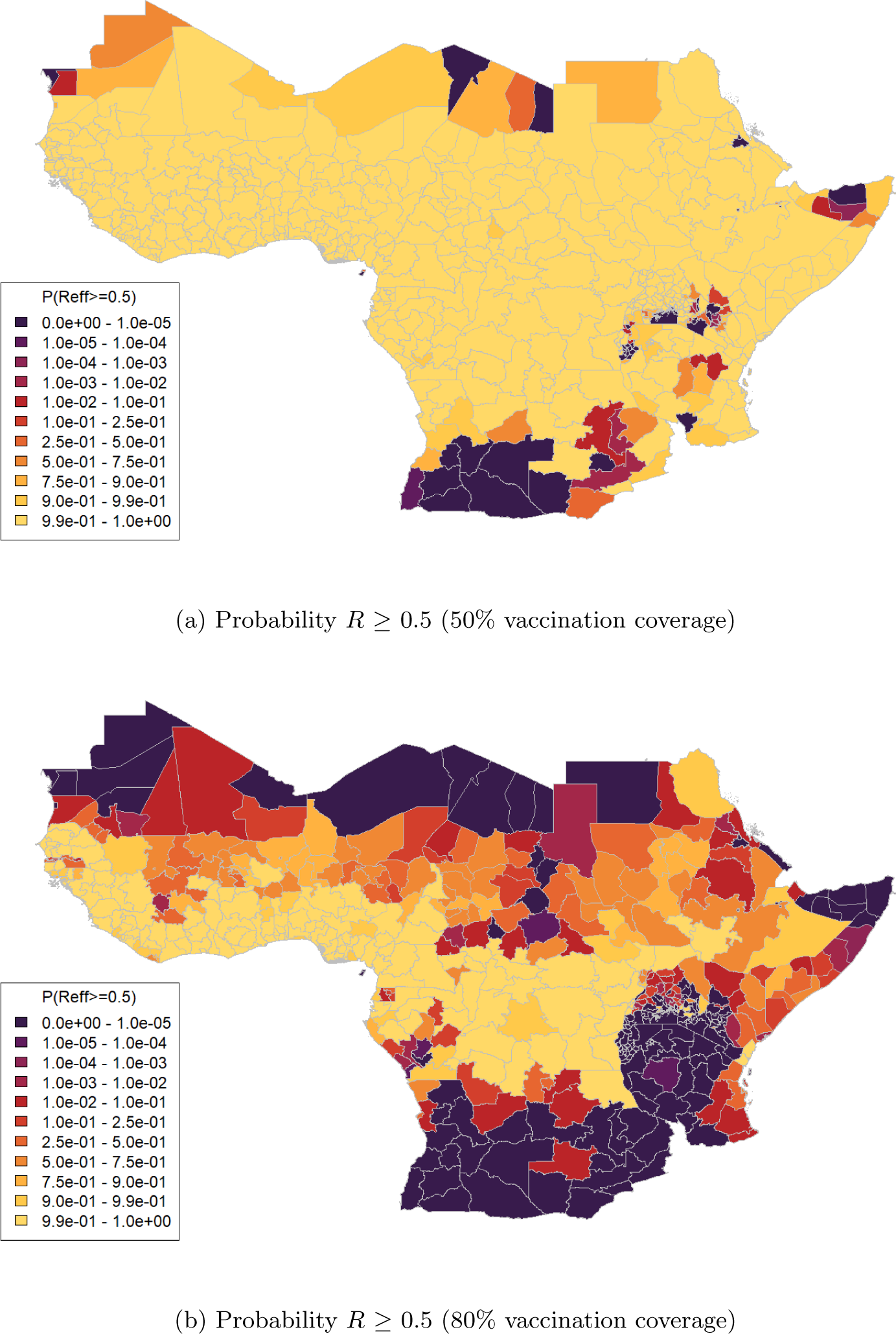
Maps of selected 1st-level sub-national administrative regions in Africa displaying (a) probability that projected *R ≥* 0.5 for vaccination coverage 50% in 1-60 year olds b) probability that projected *R ≥* 0.5 for vaccination coverage 80% in 1-60 year olds, based on distributions of values of *R*_0_ and *v_eff_* obtained from MCMC results

Additional maps are shown in the supplementary information, in section 6. These include maps for the intermediate criterion *R ≥* 0.7, and maps for scenarios in which the reported vaccination efficacy *v_eff_* was set to 97.5% (representing perfect vaccination reporting and estimated median vaccine efficacy ^25^, with all reported vaccinations assumed to be carried out successfully).

## 4 Discussion

We projected the impact of the EYE Strategy vaccination targets on preventing self-perpetuating outbreaks through reducing the effective reproduction number *R* using a novel model of YF transmission. This model, in contrast to other recent studies ^30^, captures outbreak dynamics and was estimated from available data such as serological studies and case notifications. We find a high probability that the value of *R*_0_ *≥* 1 across large parts of the regions of Africa where YF is endemic. When we consider the EYE strategy targets and the effective reproduction number, it is probable that *R* can be brought below 1 through reaching 50% vaccination coverage in target populations. However, this should be treated with caution as seasonality, climate change, and other factors like land-use change, may mean a changing landscape of YF outbreak risk.

Seasonal variation, extreme weather events such as flooding, climate change and/or other human-driven changes over time (e.g. major changes in land use) may cause substantial inter- and intra-annual fluctuations in transmission. To account for this, we assessed the EYE Strategy target using more stringent criteria, namely its ability to reduce *R* below 0.7 or 0.5. It was found that the lower EYE Strategy vaccination target was unable to achieve this across the majority of Africa (Figure 7). Increasing vaccination coverage to 60% or 80% reduces the number of at-risk regions but there are some regions that may be vulnerable if transmission variability is high.

These results suggest that the EYE Strategy targets are sufficient to eliminate outbreaks in most regions in Africa. However, regions with high seasonal variation or areas where there is a large proportion of vaccine misclassification may be at risk. Vaccination targets may therefore need to be reviewed in future, e.g. via increasing the coverage in the target age group and/or via expanding the target age range. The effort required to boost population level vaccination coverage will vary by region and existing vaccination levels ^18^.

In our study, we estimate a measure of vaccination effectiveness which encompasses both vaccine efficacy and misclassification/misreporting of vaccination coverage. We used estimates of vaccine efficacy ^25^ as our prior information and our posterior estimates suggest a degree of vaccine coverage misreporting and/or a lower vaccine efficacy than previously estimated. Regarding efficacy, it has been suggested that the long-term immunity in children may be lower than in vaccinated adults; Domingo et al. found substantial decreases in seropositivity in children following immunisation suggesting that one dose may not be sufficient for endemic countries ^31^. Regarding misclassification, a recent study estimated the sensitivity and specificity of vaccination misclassification and found both to be approximately 70-75% which may suggest that our lower estimate of vaccine efficacy is partly accounting for misclassification in vaccination status ^26^. Thus, whilst the individual study estimate of vaccine efficacy may be higher, our estimate should be considered a composite measure of vaccine efficacy and implementation.

A key challenge to the modelling work reported here was the sparse and limited nature of the data available to use to estimate model parameters. Serological surveys provide only ”snapshots” of certain regions in certain years, whereas annual reported case/death data is only available nationally in a subset of countries and may be affected by under-reporting. As more and varied data continues to become available, estimates will be able to be refined and uncertainty reduced.

This work includes some limitations. An important internal limitation is the assumption that values of the spillover force of infection, basic reproduction number and reporting probabilities are static over the course of the modelled time periods. The static values are assumed to represent a year-round average of seasonally varying values. There is insufficient information available to accurately model seasonal transmission variation across many different regions. Furthermore, we assume reporting probabilities do not vary spatially where, in reality, they will be affected by different processes and population densities. We also assume disease progression and transmission is independent of age and gender, yet it has been noted ^32^ that working age males are at a higher risk of YF infection than other population groups. Human movement is not explicitly incorporated and so the propagation of risk through human mobility will not be captured here, although the spillover force of infection will incorporate some measure of external risk. In addition, we assume lifelong immunity to yellow fever; however, as mentioned, some recent results suggest that immunity may wane following childhood vaccination ^31;33;34^. The inclusion of waning immunity in the model could potentially reduce overall population immunity (supporting the use of more stringent requirements for vaccination coverage to reduce *R* values).

## 5 Conclusions

Using a dynamic model of yellow fever transmission estimated from available epidemiological data, we have evaluated the potential for the EYE Strategy vaccination targets to eliminate yellow fever outbreaks driven by human-to-human transmission. The results indicate that the EYE Strategy targets are sufficient to eliminate large outbreaks based on an average annual basic reproduction number, but that for some high burden regions, they may not be sufficient if seasonal or climatic variation of the basic reproduction number are taken into account. These results suggest that the interim 2022 EYE Strategy target of 50% coverage may not be sufficient for the complete elimination of large yellow fever outbreaks, particularly in areas with high seasonal range, but that the 2026 EYE Strategy targets of 60-80% coverage significantly reduce outbreak risk.

## Data Availability

Data on reported yellow fever case and death numbers used in this study are publicly available. Seroprevalence data used in this study is partially available in referenced publications, but this study used raw data which is confidential.
All code used for the numerical modelling work in this study will be made available on request. The software package containing the model code and functions used to handle input and output data is publicly available with documentation at https://github.com/mrc-ide/YEP.

## 6 Declarations

### 6.2 Data availability

Population and vaccination data used in this study is publicly available ^15;18^. Yellow fever seroprevalence data used in this study is taken from previously published studies (see supplementary information for details); we used raw data from these studies which is confidential. Yellow fever annual case data used in this study is publicly available ^35^.

### 6.3 Code availability

A custom-created R package (YEP, Yellow Fever Epidemic Prevention - https://github.com/mrc-ide/ YEP/) was used for data analysis; this package is publicly available. It makes use of other publicly available custom R packages including odin.dust (https://github.com/mrc-ide/odin.dust). Specific code used for simulations and data processing in this paper can be made available on request.

