## Supplementary Information for "Assessing yellow fever outbreak potential and implications for vaccine strategy"

### 1 Introduction

We describe our model of yellow fever infection incorporating vaccination, population and transmission dynamics, incorporating both infection due to spillover from the sylvatic (non-human primate) reservoir and infection due to human-to-human transmission. We further describe how epidemiological parameter values were estimated within a Bayesian framework based on observed data.

The odin.dust package[1] was used to implement the SEIRV model; this is a domain specific language based on R which is automatically compiled to C++ for improved computation speed. Additional functions for input data preparation, output data processing, parameter estimation and other tasks were created in R. A downloadable R package containing updated versions of the relevant functions is available at https://mrc-ide.github.io/YEP.

### 2 SEIRV model

#### 2.1 Overview

We used a compartmental model in which the population of a region (formed of one or more sub-national regions defined by version 3.6 of the Database of Global Administrative Areas[2]) was divided into susceptible (S), exposed (E), infectious (I), recovered (R) and vaccinated (V) groups. The population was further divided by age into annual subgroups up to age 100. The key variable parameters were:

- The force of infection due to spillover from the sylvatic reservoir and/or importation of infections from outside a region, $\lambda_S$

- The basic reproduction number for mosquito-mediated human-to-human transmission, $R_0$
- The reported vaccination effectiveness (i.e. the proportion of reported vaccinations which result in an individual becoming protected), $v_{eff}$
- The probability of a severe infection being reported and confirmed via lab testing as a case, $P_{R,S}$
- The probability of a fatal infection being reported and confirmed via lab testing as a case, $P_{R,D}$

Note that it was assumed that mild or asymptomatic infections are not reported as cases.
Fixed parameters included:

- The yellow fever incubation time in mosquitoes $t_{INC}$ (set to 5 days)
- The latent period in humans between exposure and the onset of infectiousness $t_L$ (set to 5 days)
- The period of infectiousness in humans $t_{INF}$ (set to 5 days)
- The proportion of infections which cause severe symptoms $P_S$ (set to 12%)
- The proportion of severe infections which lead to death $P_{S,D}$ (set to 39%)
- The time increment $dt$ between each time point at which calculations are carried out (set to 5 days)

##### 2.2 Initial conditions

The initial population in each age group was derived from available population data[3]. The proportion initially vaccinated in each age group was derived from available data on estimated population immunity due to vaccination[4]. The proportion recovered in each age group was set to give herd immunity within that age group at the start of the modelled period (in 1940, for the work described here), reflecting a situation prior to widespread vaccination where yellow fever was endemic. Immunity was set as a function of age, using equation 1, where $F_R(A)$ and $F_V(A)$ are the fractions of the population in age group A who have infection-induced and vaccine-induced immunity respectively and $\lambda_{est}$ is an estimated force of infection calculated by non-linear fitting to give an overall immunity level across all ages of 1-1/$R_0$.

$$F_R(A) = (1 - exp(\lambda_{est}(A + 0.5))) - F_V(A) \quad (1)$$

##### 2.3 Calculation of parameters from environmental covariates

The epidemiological parameters $\lambda_S$ and $R_0$ were calculated from environmental covariates $C_{ENV}$ including population, non-human primate density and temperature suitability[5] (see section 2.2.2 in the main text). The calculation was carried out using linear formulae (Equations 2) with coefficients $M_{\lambda,I}$ and $M_{R,I}$ for each covariate $C_{ENV,I}$. It should be noted that the dependence on temperature is nonlinear as it is assumed that $\lambda_S$ and $R_0$ are linear functions of the temperature suitability index, a nonlinear function of mean temperature estimated previously[5].

$$\lambda_S = \sum (C_{ENV,I} M_{\lambda,I}),$$
$$R_0 = \sum (C_{ENV,I} M_{R,I}). \tag{2}$$

The estimation of the coefficients $M_{\lambda,I}$ and $M_{R,I}$ using observed data is described in section 3. $\lambda_S$ values for regions in Brazil were multiplied by the adjustment factor $F_{Brazil}$ to take into account NHP surveillance (see section 2.1.1 in the main text).

#### 2.4 Calculations and outputs

The numbers of people in each subgroup $S$, $E$, $I$, $R$ and $V$ divided by age group $A$ (with each age group being 1 year wide, so that age group 1 encompasses people aged 0-12 months) were adjusted over time as shown in Equations 3 below. Note that population adjustment was not applied to groups E and I to reduce complexity, due to the small proportion of the population in groups E and I at any given time.

$$\begin{aligned}
\frac{dS(A)}{dt} &= -E_{NEW}(A) - R_{V,S}(A) + \Delta P_S(A), \\
\frac{dE(A)}{dt} &= E_{NEW}(A) - \frac{E(A)}{t_{INC} + t_L}, \\
\frac{dI(A)}{dt} &= \frac{E(A)}{t_{INC} + t_L} - \frac{I(A)}{t_{INF}}, \\
\frac{dR(A)}{dt} &= \frac{I(A)}{t_{INF}} - R_{V,R}(A) + \Delta P_R(A), \\
\frac{dV(A)}{dt} &= R_{V,S}(A) + R_{V,R}(A) + \Delta P_V(A).
\end{aligned} \tag{3}$$

The number of new infections $E_{NEW}$ of susceptible people was calculated using a total force of infection $\lambda_{TOT}$ calculated from $\lambda_S$ and $R_0$ (Equation 4). New infections were drawn from a binomial distribution, with the number of susceptible people in an age group as the number of tests and the force of infection as the probability of "success" (Equation 5).

$$\lambda_{TOT} = \lambda_S + \frac{R_0}{t_{INF}} \frac{\sum (I)}{P}, \tag{4}$$

$$E_{NEW}(A) = \binom{S(A)}{\lambda_{TOT}}. \tag{5}$$

Changes in $S$, $R$ and $V$ due to population ageing/death and new births ($\Delta P_S$, $\Delta P_R$, $\Delta P_V$) were calculated using Equations 6. $F_S$, $F_R$ and $F_V$ denote the proportion of the total population in a given age group who are in group $S$, $R$ and $V$. Note that YF deaths are assumed to be subsumed into overall population dynamics; mortality due to YF was not explicitly included.

$$\begin{aligned}
\Delta P_S(A=1) &= \Delta P_1(A=1) - \Delta P_2(A=1)F_S(A=1),\\
\Delta P_S(A>1) &= \Delta P_1(A)F_S(i-1) - \Delta P_2(A)F_S(A),\\
\Delta P_R(A=1) &= -\Delta P_2(A)F_R(A),\\
\Delta P_R(A>1) &= \Delta P_1(A)F_R(i-1) - \Delta P_2(A)F_R(A),\\
\Delta P_V(A=1) &= -\Delta P_2(A)F_V(A),\\
\Delta P_V(A>1) &= \Delta P_1(A)F_V(i-1) - \Delta P_2(A)F_V(A).
\end{aligned} \tag{6}$$

The numbers of people granted immunity via vaccination from subgroups $S$ and $R$, $R_{V,S}$ and $R_{V,R}$ were calculated as shown below (equation 7), based on the fractional vaccination rate (proportion of the population in a given age group vaccinated in a given year) $R_V$ and the reported vaccination effectiveness $v_{eff}$. $R_V$ in turn was calculated for each age group A and year Y from separately calculated estimated population immunity ($I_V(A,Y)$) due to vaccination[4] (equation 8). $F_{S,NV}$ and $F_{R,NV}$ denote the susceptible and recovered portions of the unvaccinated population ($S$+$R$). Note that it was assumed here that susceptible and recovered individuals have an equal chance of being vaccinated. Potential "double vaccination" of already vaccinated individuals was already taken into account in the separately calculated vaccination data.

$$\begin{aligned}
R_{V,S}(A) &= R_V(A)F_{S,NV}(A)v_{eff},\\
R_{V,R}(A) &= R_V(A)F_{R,NV}(A)v_{eff}.
\end{aligned} \tag{7}$$

$$R_V(A,Y) = P(I_V(A+1,Y+1) - I_V(A,Y)) \tag{8}$$

The model outputs the values of $S$, $E$, $I$, $R$, and $V$ for each age group at each time interval, along with the number of newly infectious individuals $C$ (equal to the number transferred from $E$ to $I$) across all age groups. These were used to calculate the following types of output data, corresponding to the types of real-world epidemiological data used for parameter estimation:

- Seroprevalence values for one or more age ranges averaged over a given year,
- Annual reported and confirmed severe and fatal cases (calculated using $P_S$, $P_{R,S}$, $P_{S,D}$ and $P_{R,D}$) for a given year.

### 3 Parameter estimation methods

Estimation was conducted using both the stochastic model and its deterministic expectation; the results were qualitatively similar and so the deterministic expectation was used for the final, real-world data estimation.

#### 3.1 Parameter set

The set of parameters estimated via the methods described below consists of the coefficients of the environmental covariates $M_{\lambda,I}$ and $M_{R0,I}$ along with the reporting probabilities $P_{R,S}$ and $P_{R,D}$, the reported vaccination effectiveness $v_{eff}$ and, where relevant, the Brazil surveillance factor $F_{Brazil}$ (see section 2.1.2 in the main text).

To ease chain mixing, a log transform was used for all parameters.

##### 3.2 Prior distribution

We assumed the prior distribution for each model parameter was a truncated normal distribution defined by a mean value $mu$ and standard deviation $sigma$ with permitted maximum and minimum values.

A logarithmic truncated normal distribution was used in the case of the coefficients of the environmental covariates $M_{\lambda,I}$ and $M_{R0,I}$, and a non-logarithmic distribution in the case of the additional parameters $v_{eff}$, $P_{R,S}$, $P_{R,D}$ and $F_{Brazil}$. Values of $M_{\lambda,I}$ and $M_{R0,I}$ were restricted only to positive values. The values of $v_{eff}$, $P_{R,S}$, $P_{R,D}$ and $F_{Brazil}$ were further restricted to values between 0 and 1.

An additional prior was applied to values of $R_0$ calculated from the environmental covariates; this was calculated using a truncated normal probability formula with mean 4.8 and standard deviation 1 informed by Liu and Rocklov 2020[6].

##### 3.3 Likelihood calculation

To assess a given set of model parameters, we calculate the likelihood of observing a complete set of observed serological and/or annual case data based on the simulated data generated using the parameter set. This was calculated across all the elements of the dataset, by adding together the natural logarithm of the likelihood of observing each individual element to give the total logarithmic likelihood.

The likelihood of observing one element of serological data taken from a survey (a number of positive tests $x$ for a total number of individuals tested $t$) was calculated using equation 9, where $D_{binom}$ is a binomial probability density function and $P_{sero}$ is the seroprevalence in the relevant region and age group, calculated from SEIRV data output from the model using the proportion of individuals in the survey who were known to be unvaccinated. If all tested individuals are unvaccinated, $P_{sero}$ is given by the ratio $R/(S + E + I + R)$, but the calculation becomes more complicated if some individuals' vaccination status is unknown[7]. Equation 10 shows how $P_{sero}$ was calculated in this case. $F_{V,U}$ is the proportion of the group tested in the survey for whom vaccination status was unknown, and therefore a positive test result might be caused by the individual being vaccinated.

$$LogLike_{sero} = log(D_{binom}(positives = x, samples = t, prob = P_{sero})). \quad (9)$$

$$P_{sero} = F_{V,U}((R + V)/(S + E + I + R + V)) + (1 - F_{V,U})(R/(S + E + I + R)). \quad (10)$$

The likelihood of observing one element of annual severe or fatal case data (a number of recorded severe or fatal cases $C_{obs}$) was calculated using equation 11, where $D_{nbinom}$ is a negative binomial probability density function and $C_{model}$ is the calculated number of cases.

$$LogLike_{case} = log(D_{nbinom}(x = C_{obs}, \mu = C_{model})). \quad (11)$$

The total logarithmic posterior probability of observing the data for a given parameter set was calculated by adding the total logarithmic likelihood ($LogLike_{sero}$+$LogLike_{case}$ to the total logarithmic prior probability $p_prior$.

##### 3.4 Markov Chain Monte Carlo sampling

Adaptive Markov chain Monte Carlo (MCMC) sampling was used to estimate the posterior distribution of the parameter values within a Bayesian framework. At each iteration of the Markov chain, a new proposed set of model parameters $C_{prop}$ was generated using a random multivariate normal function (Equation 12), where the mean values $C$ are the previous parameter values and the standard deviation values $\sigma$ are calculated from the covariance $cov_{chain}$ of the chain so far (Equation 13[8]).

$$C_{prop} = R_{mvnorm}(n = 1, \mu = C, \sigma = \sigma), \tag{12}$$

$$\sigma = \frac{(2.38^2).cov_{chain}}{n_{params}}. \tag{13}$$

For each repetition, the total posterior probability was calculated as described in section 3.3 for the proposed set of model parameters. An "acceptance probability" $p_{accept}$ was then calculated using Equation 14, where $Post_{prop}$ is the calculated logarithmic posterior probability of the proposed parameter set and $Post_{current}$ is the calculated logarithmic posterior probability of the most recent accepted parameter set (this was initially set to $-Inf$). The proposed parameter set was accepted or rejected based on this acceptance probability if a random sample from Unif(0,1) was less than $p_{accept}$.

$$p_{accept} = exp(Post_{prop} - Post_{current}). \tag{14}$$

Multiple chains were run in parallel for comparative purposes, using different starting parameter values. Outputs from one or more selected chains (those converging to the highest likelihood, with convergence assessed visually from graphs of likelihood by iteration, or by the Gelman-Rubin statistic) past the chosen burn-in points were combined into a single posterior distribution of parameter values.

### 4 Validating the parameter estimation using simulated data

#### 4.1 Simulated data generation

To test the efficacy of our MCMC methods for estimating model parameters, the approach was applied to simulated data.

The model was used to generate serological and case data for 25 regions. The modelled regions all used the same population and vaccination data, but used different values of environmental covariates $Var_{1-5}$ to produce different values of $\lambda_S$ and $R_0$ from a set of 10 environmental coefficients (5 governing $\lambda_S$ and 5 governing $R_0$, one for each covariate). Initial conditions were applied naively with immunity defined by $1 - (1/R_0)$ across all ages.

Values of the environmental coefficients were chosen randomly to give a variety of $\lambda_S$ and $R_0$ values across the 25 regions within realistic ranges (daily $\lambda_S$ varies in the approximate range $10^{-8}$ to $10^{-6}$, while $R_0$ varies from around 0 to 2.5, as shown in 3). The values of $P_{R,S}$ and $P_{R,D}$ were set at 10% and 20% respectively. Reported vaccination effectiveness $v_{eff}$ was fixed at 100% throughout, and $F_{Brazil}$ was not used.

Serological and case data was generated for 25 regions for the years 1980-2000. This represents more data than was available for any real-world region; as described in section 4.2, a selection of this data was used for estimation, to represent the sparseness of real-world data.

##### 4.2 Parameter estimation results from simulated data

The model was estimated from the the simulated data (see section 4.1). To account for the sparseness of real-world data, only the following simulated data was used:

- Serological data from regions 1-15 in the year 1985,
- Annual observed severe and fatal case data from regions 17-25 in the period 1990-1999.

Latin hypercube sampling[9] was used to set the initial values of the environmental coefficients governing $\lambda_S$ and $R_0$ as well as of the reporting probabilities for severe and fatal cases, by sampling from a designated parameter space and calculating the likelihood obtained for each set of parameter values. The 4 parameter sets giving the highest likelihood were used as inputs for 4 Markov chains. Each chain was run for 100,000 iterations with 5 stochastic repetitions per iteration.

Some settings for estimation from simulated data were different from those described in sections 3.2 and 5. Reported vaccination effectiveness $v_{eff}$ was fixed at 100% in simulated data generation, as noted above, and was therefore not estimated as a variable parameter, and $F_{Brazil}$ was fixed due to not being required. The total number of parameters to be estimated using MCMC was therefore 12. No prior was applied to calculated values of $R_0$, and priors were applied to the logarithmic rather than actual values of $P_{R,S}$ and $P_{R,D}$. The logarithmic mean prior applied to the values of $M_{\lambda,I}$, $M_{R0,I}$, $P_{R,S}$ and $P_{R,D}$ was 0 with standard deviation 30 in all cases (a very loose prior allowing for free exploration of the parameter space). The only restriction on maximum and minimum values of the parameters was the restriction of $P_{R,S}$ and $P_{R,D}$ to values between 0.01 and 1.

Figure 1 shows how calculated posterior probability varied with successive MCMC iterations for the 4 Markov chains using the 4 different starting parameter sets.

The 4 displayed chains converge on the same range of likelihood values, with the Gelman-Rubin convergence parameter[10] having a value of 1.24 (below the suggested threshold for convergence of 1.3). Parameter values were therefore combined from all 4 displayed chains (using values from approximately 50,000 to 100,000 iterations, i.e. the second half of each chain) to confirm that the MCMC approach correctly estimates the epidemiological parameters used to create the "observed" data. Values of $\lambda_S$ and $R_0$ were calculated from the environmental coefficient parameters as described in section 2.3.

Violin plots of obtained values of the coefficients used to generate $\lambda_S$ and $R_0$ values from the values of the environmental covariates are shown in Figures 2a-b in black, along with the correct original values in red. Figure 2c shows a violin plot of values of $P_{R,S}$ and $P_{R,D}$ obtained in the same way, again compared with the correct original values. It can be seen that the MCMC approach produces broad ranges of parameter output values encompassing the correct values.

Violin plots of $\lambda_S$ and $R_0$ values for each simulated region (obtained via calculation using the environmental coefficients shown in figures 2a-b) are shown in figures 3a-b for each region in black, along with the correct original values (obtained from the real coefficient values) in red. The spread of values was lower than for the enviromental coefficients, giving a closer match These results show that the MCMC approach produces distributions of values which can be used to duplicate the epidemiological parameters underlying the data.

Figures 4a-c below illustrate the matching of the simulated serological and annual severe/fatal case data to values generated using parameters output from the chain. 1000 sets of parameter values taken from regular intervals from the dataset taken from the second half of each of the 4 chains were used to generate new data, with 95% (blue) and 50% (green) bounds of the resulting distributions of values shown on the graphs below.

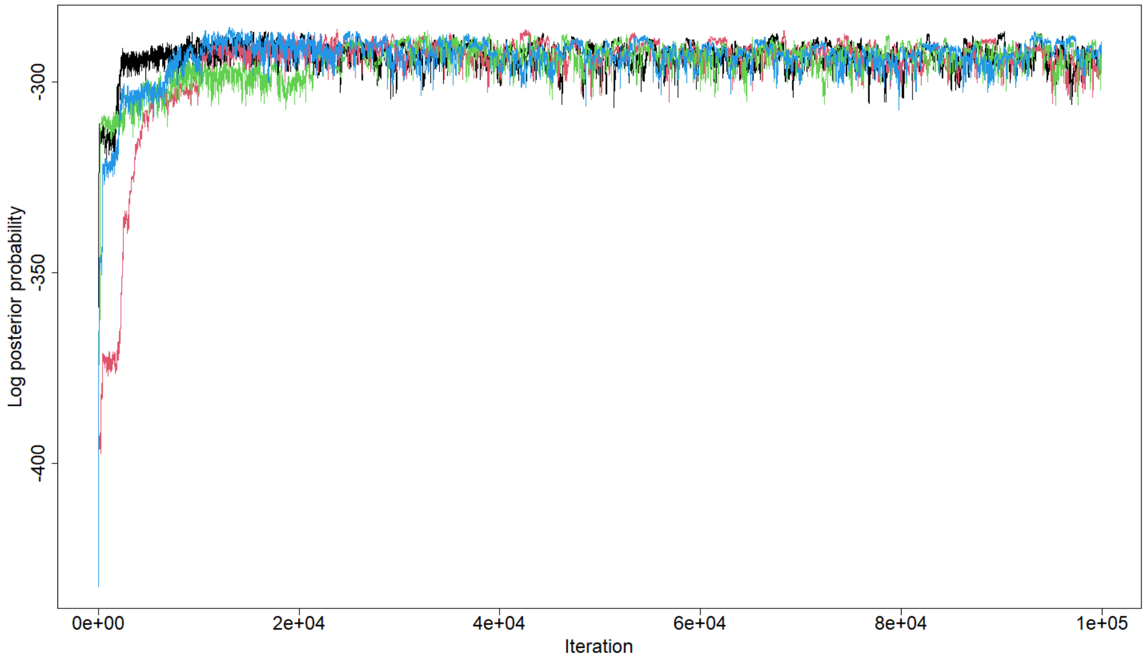


Figure 1: Calculated posterior probability by iteration for 4 Markov chains based on selected simulated serological and annual reported severe and fatal case data.

### 5 Parameter estimation based on real data

#### 5.1 Data

Serology data to be used for parameter estimation was compiled from 17 cross-sectional surveys [11–19] carried out between 1985 and 2019 in a total of 13 countries (Central African Republic, Cameroon, Democratic Republic of the Congo, Republic of the Congo, Ethiopia, Kenya, Nigeria, Rwanda, Sudan, South Sudan, Chad, Uganda, Zambia) in Africa. The data in each survey was gathered in one or more 1st-level sub-national regions (from a single region in some surveys to several groups of regions or an entire country in others), which was taken into account in matching model results with it. This data set matched that used for previous modelling work [5;20;21] with the addition of one recent survey [18]. In the majority of surveys, all tested individuals were unvaccinated, but in a small number, some individuals' status was unknown. This can affect the interpretation of the data [7] and the proportion of vaccinated individuals was therefore estimated within the modelling process (see section 3.3).

Public data compiled by the Pan American Health Organization [22] was used as the source of annual reported numbers of cases and deaths in South American countries. Similar data is available for other countries [23]; selected data for 11 African countries was used for validation (see section 5.3). Data from 6 countries (Bolivia, Brazil, Colombia, Ecuador, Peru, Venezuela) was used for estimation, with these countries being chosen for the relatively high number of reported cases (≥50 per country in the period 1990-2015). Data from countries with lower case counts was omitted due to the difficulty of estimating based on very sparse data and the likelihood that some countries with low case counts experience yellow fever only in a few regions.

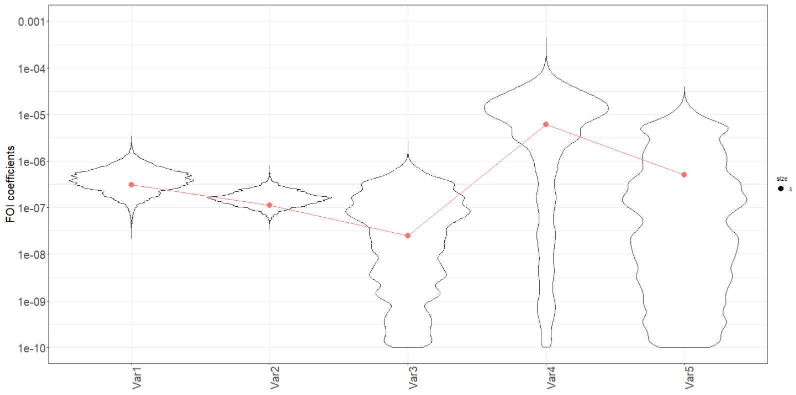


(a) Coefficients of environmental covariates used to calculate $\lambda_S$

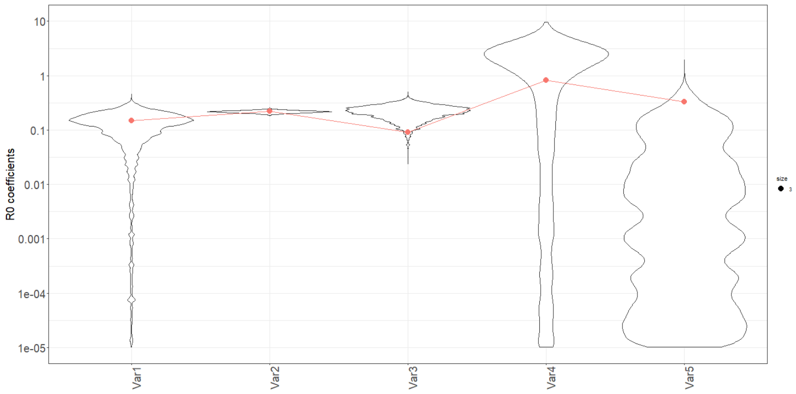


(b) Coefficients used to calculate $R_0$

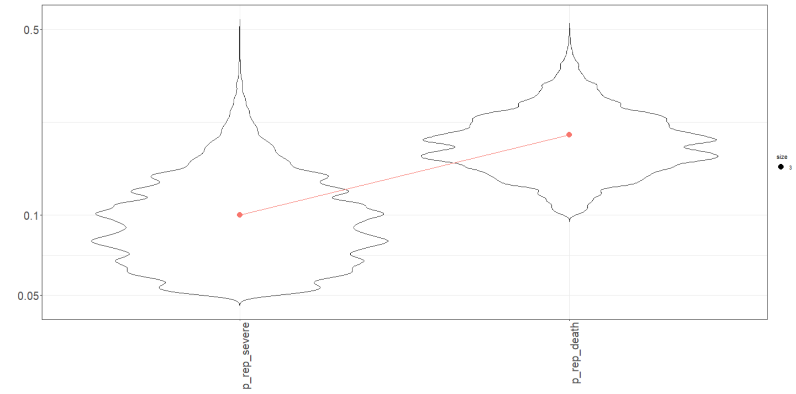


(c) Severe and fatal case reporting probabilities

Figure 2: Violin plots of parameter values (black) obtained from Markov chains shown in Figure 1 (iterations 50,000-100,000), compared with values used to create simulated data (red): a) coefficients of environmental covariates used to calculate $\lambda_S$ b) coefficients used to calculate $R_0$ c) severe and fatal case reporting probabilities. The y-axes in plots (a) and (b) are truncated to remove coefficient values very close to zero.

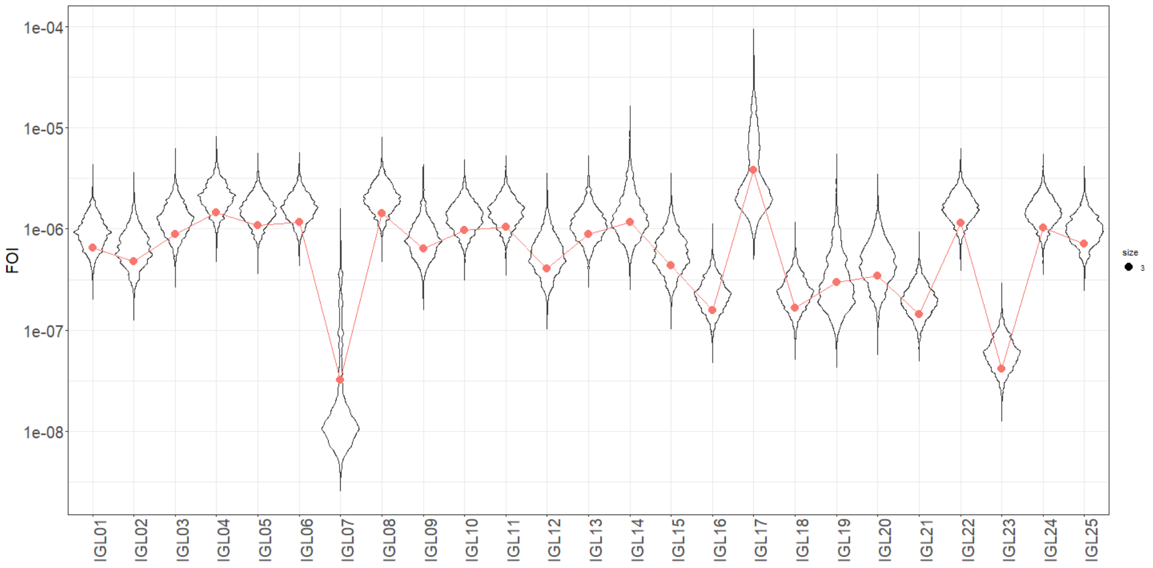


(a) $\lambda_S$ values by region

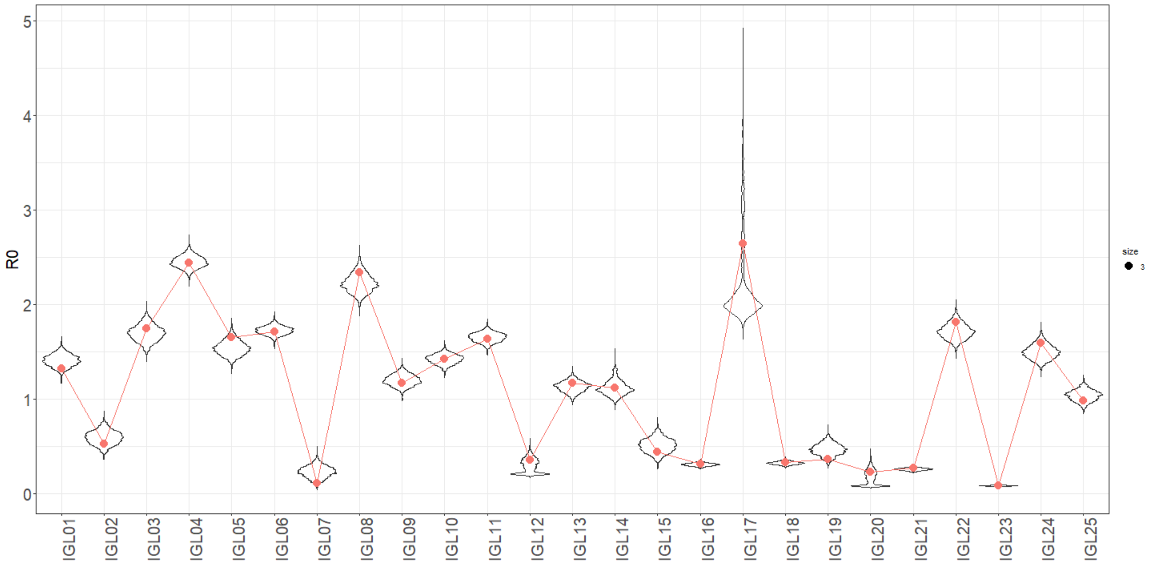


(b) $R_0$ values by region

Figure 3: Violin plots of epidemiological parameter values (black) obtained from Markov chains shown in Figure 1 (iterations 50,000-100,000), compared with values used to create simulated data (red): a) $\lambda_S$ values by region b) $R_0$ values by region.

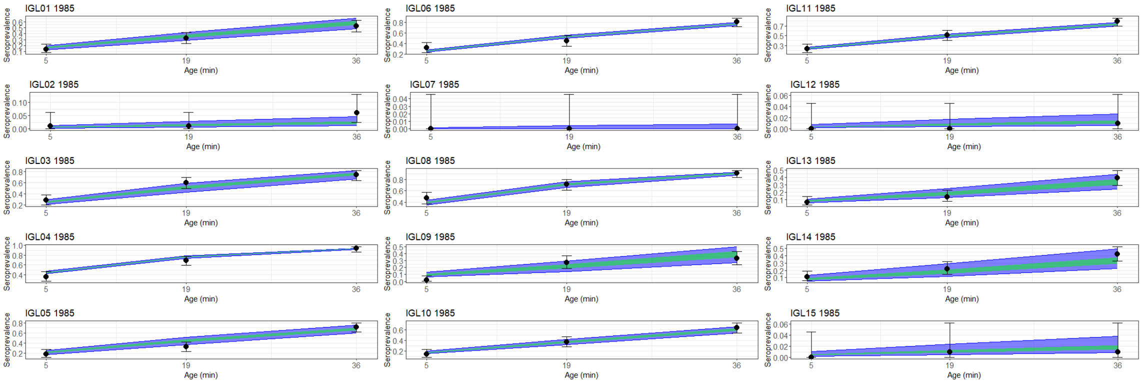


(a) Serological data

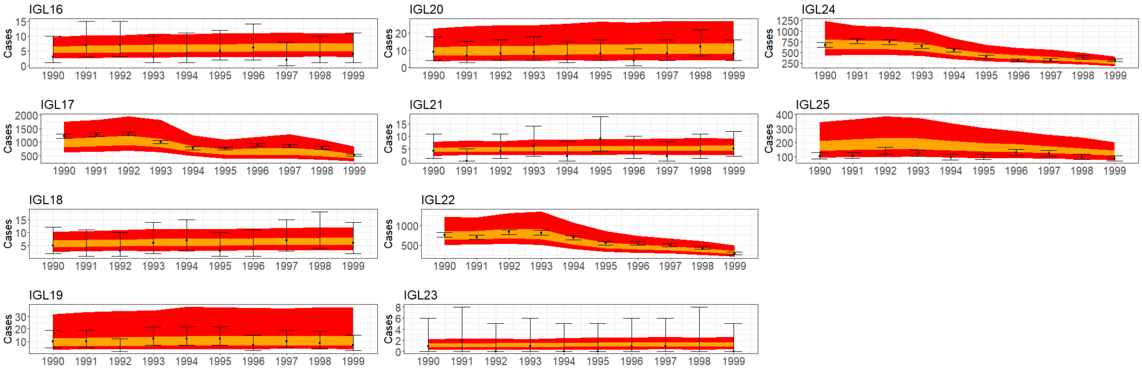


(b) Reported cases

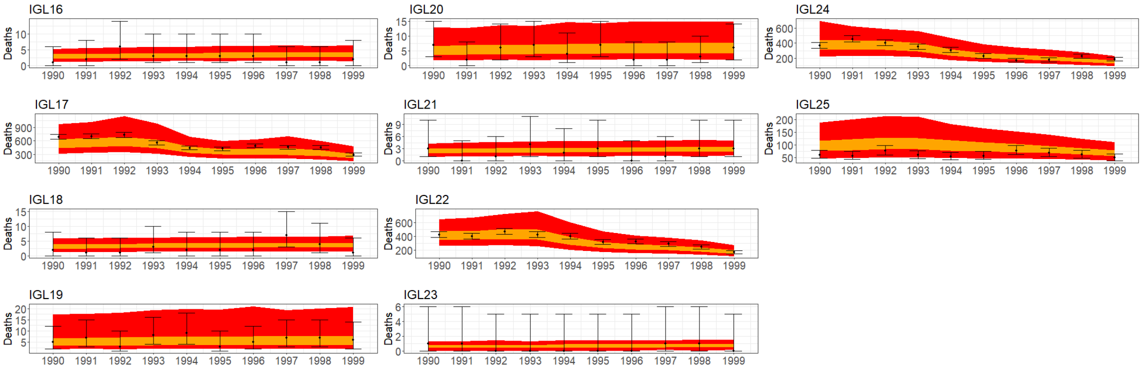


(c) Reported deaths

Figure 4: Selected simulated data (black) for regions used for MCMC parameter estimation, compared with distribution of values generated by 1000 parameter value sets drawn from the overall distribution (blue = 95% confidence interval, green = 50% confidence interval. a) Serological survey data) b) Annual reported case data c) Annual reported death data

| Parameter | Median | CI (lower) | CI (upper) | Prior type | Prior mean | Prior st. dev. |
|---|---|---|---|---|---|---|
| $M_{\lambda,aegypti}$ | 3.75e-07 | 2.96e-07 | 4.58e-7 | Log | 0 | 30 |
| $M_{\lambda,LC10}$ | 5.44e-14 | 1.13e-21 | 1.55e-08 | Log | 0 | 30 |
| $M_{\lambda,logpop}$ | 3.35e-14 | 1.54e-29 | 1.32e-09 | Log | 0 | 30 |
| $M_{\lambda,MIR}$ | 2.06e-13 | 3.20e-29 | 2.51e-08 | Log | 0 | 30 |
| $M_{\lambda,NHP}$ | 4.75e-12 | 2.51e-25 | 4.72e-09 | Log | 0 | 30 |
| $M_{\lambda,temp-suit}$ | 8.61e-09 | 6.78e-09 | 1.08e-08 | Log | 0 | 30 |
| $M_{R,aegypti}$ | 2.12e-01 | 1.87e-01 | 2.38e-01 | Log | 0 | 5 |
| $M_{R,LC10}$ | 8.92e-04 | 1.09e-05 | 1.96e-02 | Log | 0 | 5 |
| $M_{R,logpop}$ | 6.73e-02 | 5.95e-02 | 7.45e-02 | Log | 0 | 5 |
| $M_{R,MIR}$ | 6.60e-03 | 1.78e-04 | 6.06e-02 | Log | 0 | 5 |
| $M_{R,NHP}$ | 3.62e-02 | 2.95e-02 | 4.26e-02 | Log | -7 | 5 |
| $M_{R,temp-suit}$ | 7.67e-03 | 6.83e-03 | 8.24e-03 | Log | 0 | 5 |
| $v_{eff}$ | 6.08e-01 | 5.59e-01 | 6.51e-01 | Value | 0.975 | 0.05 |
| $P_{R,S}$ | 1.66e-02 | 1.45e-02 | 2.03e-02 | Value | 0.1 | 0.05 |
| $P_{R,D}$ | 2.58e-02 | 2.31e-02 | 2.94e-02 | Value | 0.1 | 0.05 |
| $F_{Brazil}$ | 7.44e-03 | 2.84e-05 | 2.56e-02 | Value | 0.1 | 0.05 |

Table 1: Table summarizing parameter output distributions and prior settings. Prior mean $v_{eff}$ value taken from Jean *etal*[24]

#### 5.2 Results

The parameters to be estimated consisted of coefficients used to calculate $\lambda_S$ and $R_0$ from 6 environmental covariates (see section 2.2.2 in the main text), along with the reported vaccination effectiveness $v_{eff}$, the probabilities of reporting of severe and fatal cases $P_{R,S}$ and $P_{R,D}$ and the Brazil adjustment factor $F_{Brazil}$. Starting parameter values for 4 Markov chains were estimated via Latin hypercube sampling and likelihood maximization, as with estimation from simulated data (section 4.2). Estimation on the real-world data was carried out with the model run deterministically, using the initial conditions described in section 2.2 (age-stratified herd immunity). Prior settings were as described in section 3.2, with the mean and standard deviation values used for the prior for each parameter summarised in Table 1 below.

Figure 5 shows how calculated likelihood varied with successive MCMC iterations for the 4 Markov chains. As with the generated data, all 4 chains converge on the same range of posterior values within the period shown; as a result, all 4 were used to generate a combined distribution of output parameter values, with the burn-in value for each chain being set at 60,000.

Figures 6a-b show the distributions of the values of the coefficients of the environmental covariates used to calculate $\lambda_S$ and $R_0$, as obtained from the post-burn-in portions of all four chains in Figure 5. The y-axes of these figures are truncated at the indicated minimum values to remove very low values effectively equivalent to zero (i.e. negligible effect of the relevant covariate on $\lambda_S$ or $R_0$ for parts of the distribution). Figure 6c shows the values of reported vaccine effectiveness, reporting/confirmation probabilities of severe and fatal cases, and Brazil adjustment factor obtained in the same way.

Table 1 summarises the median and 95% CI values for each parameter.

As discussed in section 3.1 in the main text, a distribution of 1000 values drawn from the combined posterior distributions of the 4 chains was used to calculate values for the number of expected yellow fever deaths worldwide in 2018. The figure reported in the main text (median 58,900 with a CrI of 15,100-137,500) reflects a distribution of values of severe case rate $P_S$ and severe case fatality rate

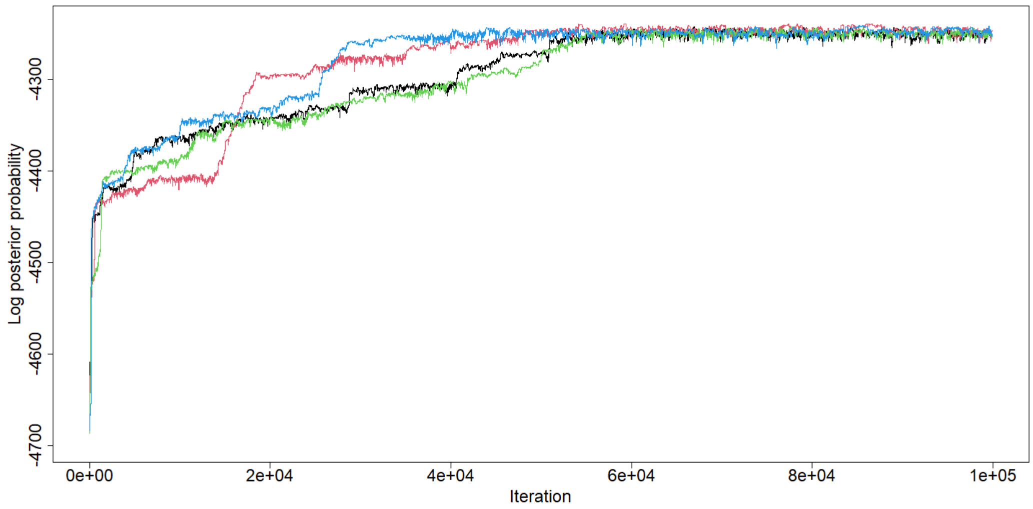


(a) All values

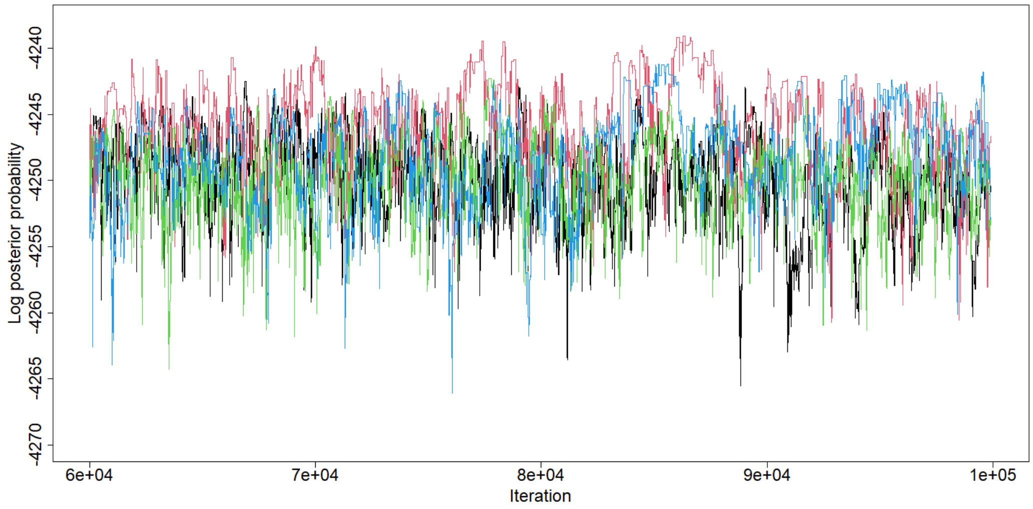


(b) Posterior distribution after burn-in

Figure 5: Calculated likelihood by iteration for 4 Markov chains based on available real-world serological and annual reported severe and fatal case data. a) All values b) Posterior distribution after burn-in.

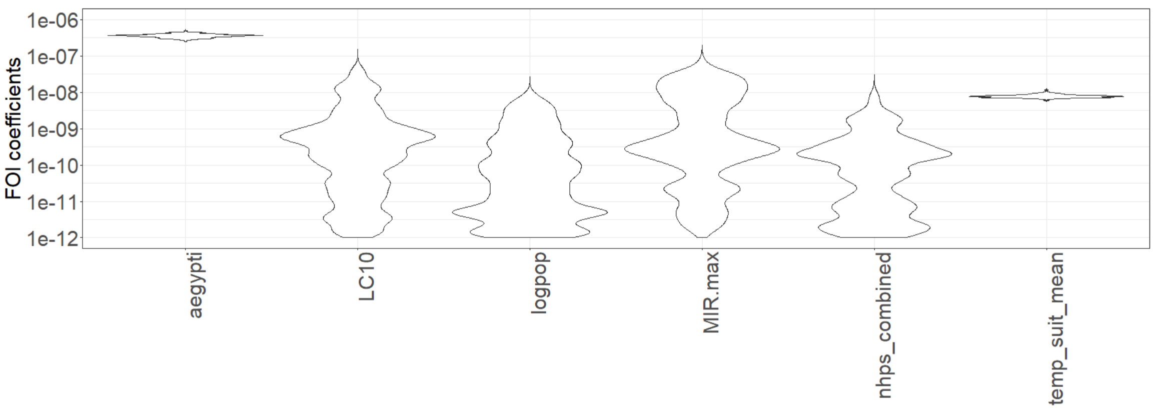


(a) Environmental coefficients- FOI

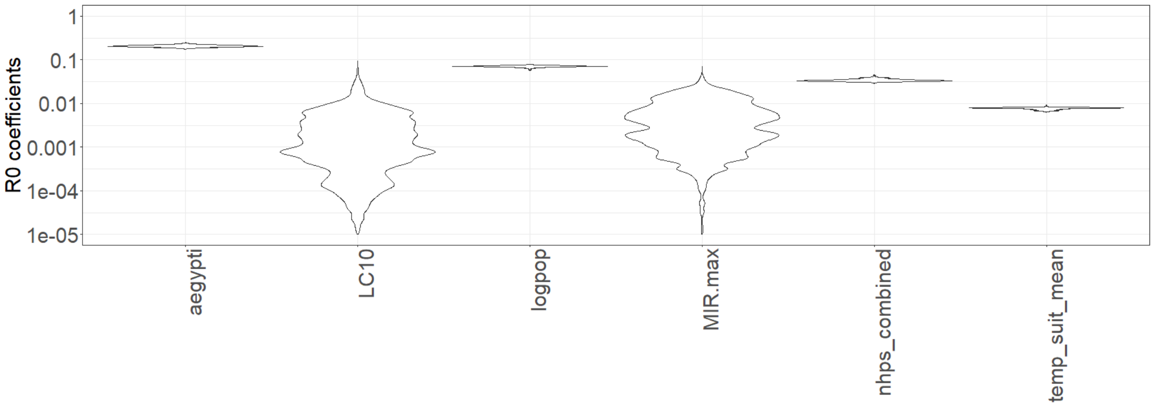


(b) Environmental coefficients- $R_0$

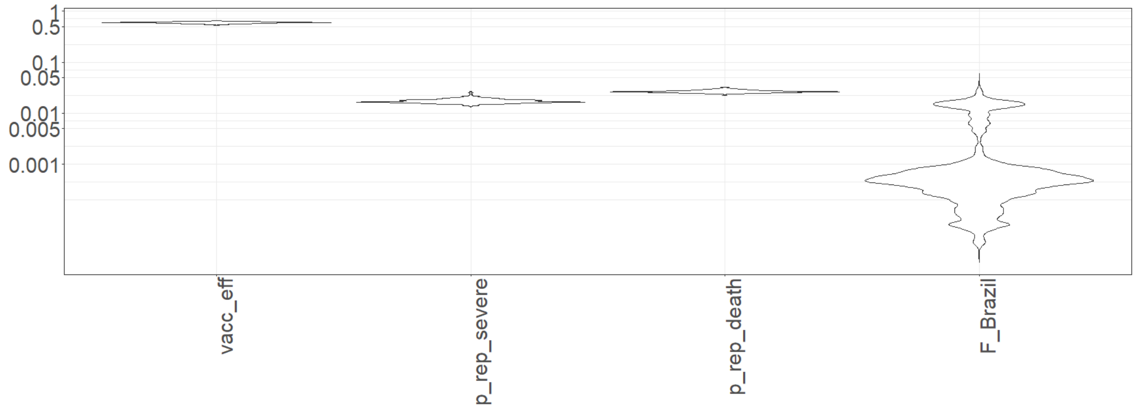


(c) Other parameters

Figure 6: Violin plots of distributions of environmental coefficient (a-b) and other parameter (c) values calculated from post-burn-in portions of Markov chains displayed in Figure 5. The y-axes in a-b are truncated at indicated minimum values to remove values close to zero.

$P_{S,D}$ taken from the source used for previous work being used for comparison[25]. If the fixed values of 12%[25] and 39%[26] used for MCMC estimation were used for these parameters, the median number of deaths was 52,800 with a 95% CrI of 48,300-58,500.

The number of deaths estimated from previous work in the literature can be adjusted to reflect the more recently estimated median severe case fatality rate[26] and compare to the above figure, giving median values of 42,000[21], 65,000[20] and 30,000[27].

##### 5.3 Validation of parameter estimate results against additional data

To validate the results of parameter estimation against other data outside that used for the estimation itself, reported case values for 11 African countries (Burkina Faso, Central African Republic, Cote d'Ivoire, Cameroon, Democratic Republic of the Congo, Ghana, Guinea, Liberia, Mali, Senegal, Sierra Leone) were simulated based on 1000 parameter sets drawn from the combined post-burn-in posterior distributions of the Markov chains. These were then compared with reported case data documented by the World Health Organization[23] (Figure 7). As with the South American case data (Figure 3 in the main text), the model produces case numbers which lie in between the upper and lower extremes of the real data, with the largest discrepancies being for individual years with very high case numbers, probably representing major outbreaks. For countries where there are substantial differences between observed and modelled values over extended periods (e.g. Cameroon and Liberia), differences in case reporting/confirmation may be partially responsible.

#### 6 Additional probability maps

As discussed in the main text (section 3.2), the probability was calculated that the basic reproduction number $R_0$ exceeded certain values in each region of interest, based on the results of the MCMC estimation of the model parameters from real data. Figure 6a in the main text shows the probability that $R_0 \geq 0.7$ across selected regions in Africa; Figure 8 displays the probability that $R_0$ exceeds 0.7 for the same regions in Africa. The probability that $R_0 \geq 0.5$ is 100% or close to it for all but a few regions (see Figure 4b in the main text).

Figure 9 shows the probability that $R_0 \geq 0.7$ across selected regions in Africa for different vaccination coverage levels, based on distributions of values of $R_0$ and $v_{eff}$ obtained from the MCMC results (compare Figure 7 in the main text).

Figure 10 shows the probability that $R_0 \geq 0.5$ across selected regions in Africa for different vaccination coverage levels, based on distributions of values of $R_0$ obtained from the MCMC results (compare Figure 7 in the main text). These maps are based on a constant reported vaccination efficacy $v_{eff}$ of 97.5%, based on estimated median vaccine efficacy[24], i.e. assuming that all reported vaccinations are carried out correctly with vaccine efficacy being the only factor limiting overall effectiveness. This naturally increases the overall effectiveness of vaccination at a given reported coverage level, such that all regions in Africa have a 100% probability of $R$ being reduced below 0.5 in this scenario if the coverage is 80% (not shown, as the map is uniform).

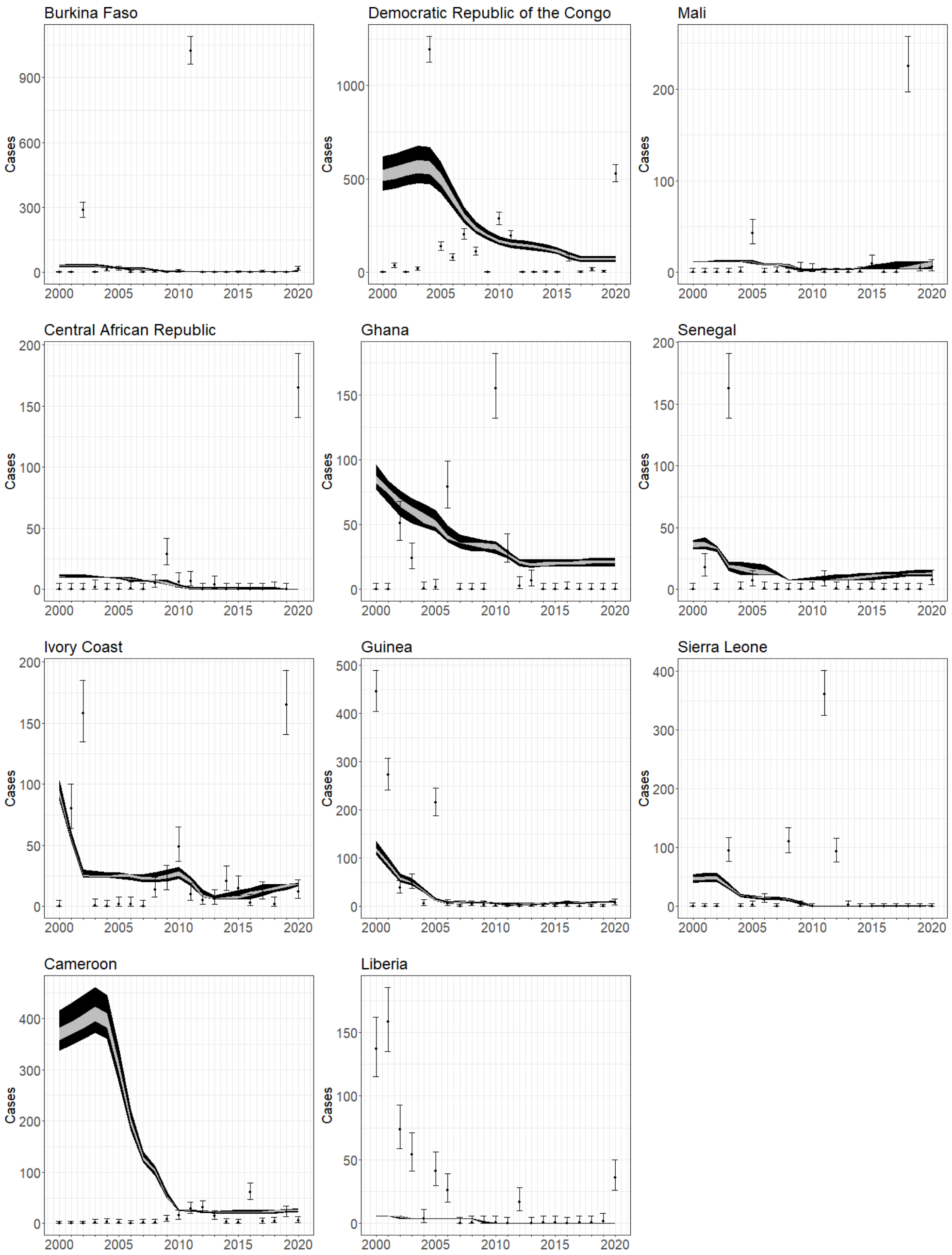


Figure 7: Graphs showing reproduction of annual countrywide reported case data for 11 African countries using 1000 parameter sets drawn from post-burn-in distribution obtained from 4 Markov chains. Points show real data (error bars obtained from binomial confidence interval calculations using the population); dark grey regions show distribution of 50% of results, light grey regions show distribution of 95% of results.

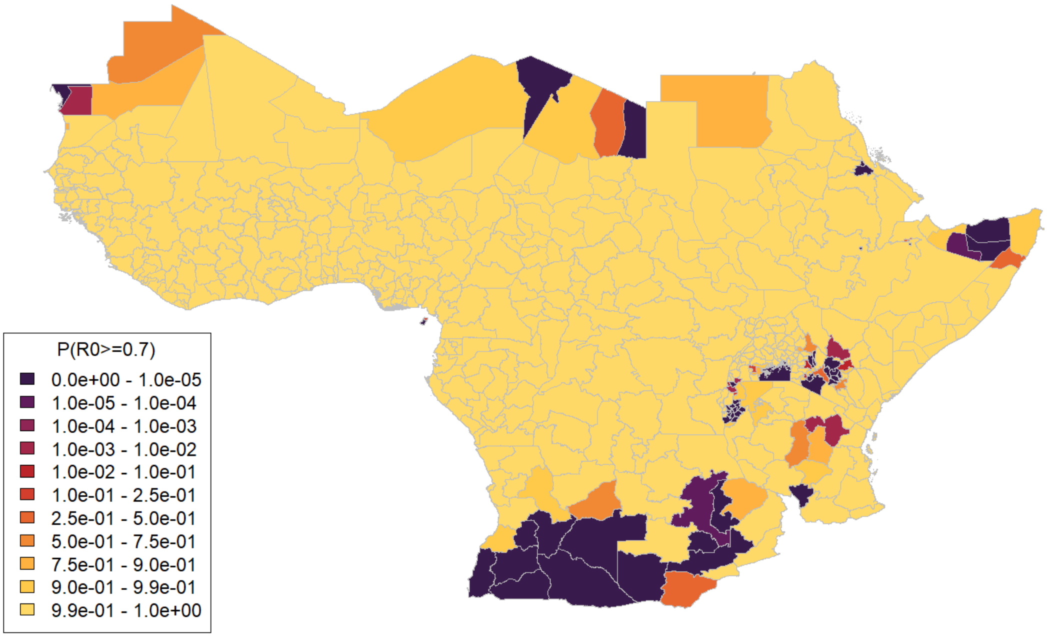


Figure 8: Map of selected 1st-level sub-national administrative regions in Africa displaying probability that $R_0$ is equal to or greater than 0.7

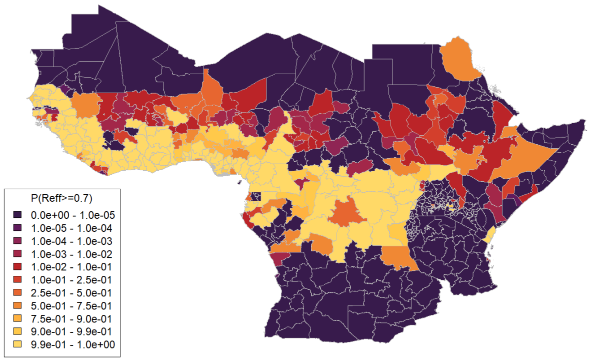


(a) Probability $R \geq 0.7$ (50% vaccination coverage)

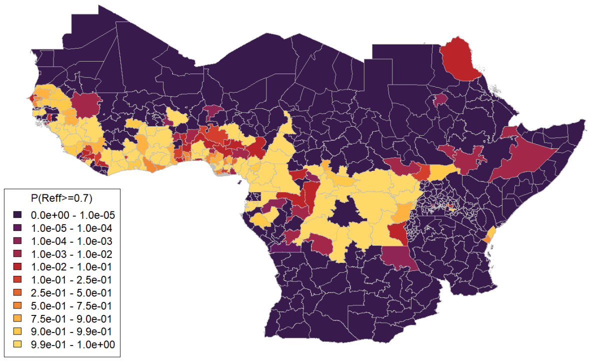


(b) Probability $R \geq 0.7$ (60% vaccination coverage)

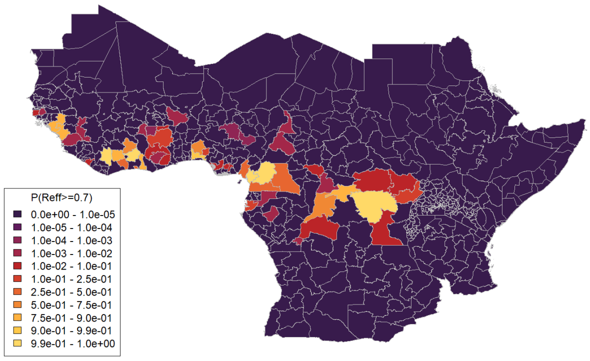


(c) Probability $R \geq 0.7$ (80% vaccination coverage)

Figure 9: Maps of selected 1st-level sub-national administrative regions in Africa displaying probability that projected $R_0 \geq 0.7$ for vaccination coverage a) 50% b) 60% c) 80% in 1-60 year olds, based on distributions of values of $R_0$ and $v_{eff}$ obtained from MCMC results.

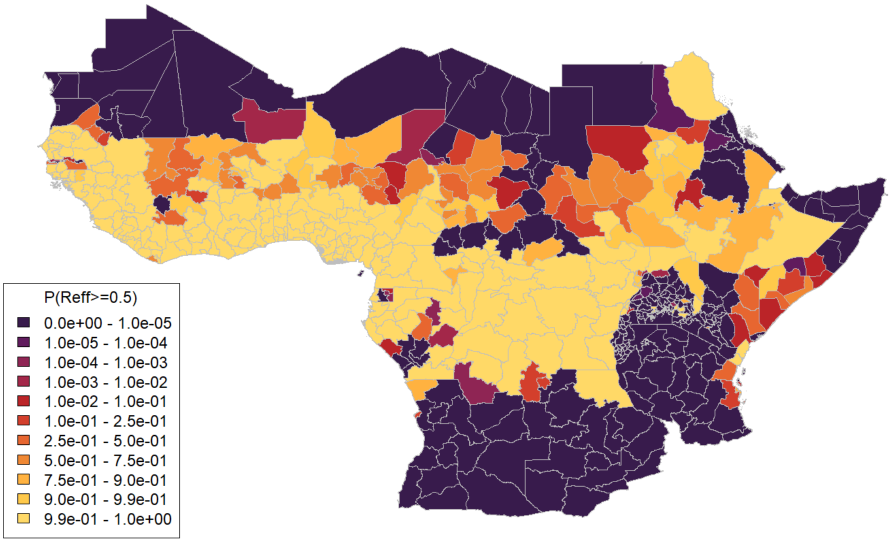


(a) Probability $R \geq 0.5$ (50% vaccination coverage, fixed $v_{eff}$)

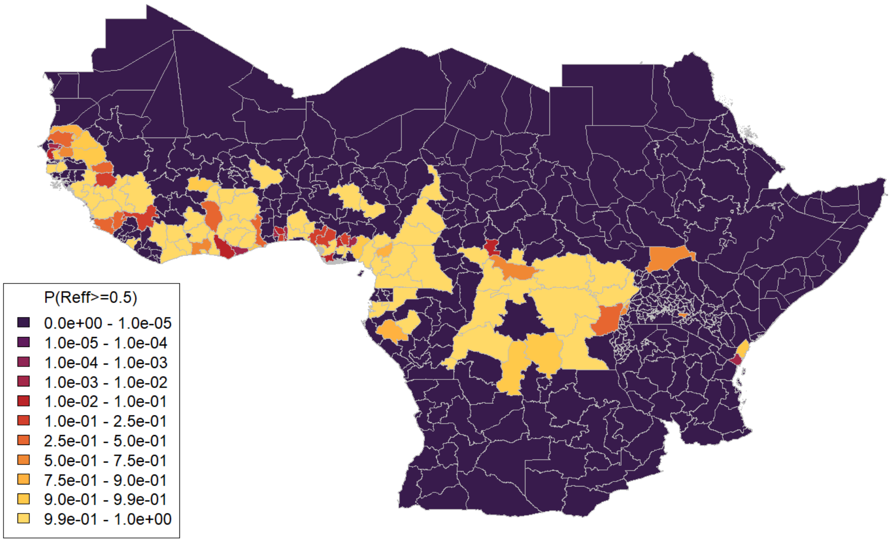


(b) Probability $R \geq 0.5$ (60% vaccination coverage, fixed $v_{eff}$)

Figure 10: Maps of selected 1st-level sub-national administrative regions in Africa displaying probability that projected $R_0 \geq 0.7$ for vaccination coverage a) 50% b) 60% in 1-60 year olds, based on distributions of values of $R_0$ obtained from MCMC results, with $v_{eff}$ set to a constant value of 97.5%.

[3] World Population Prospects; 2019.

[15] Wolfe ND, Tamoufe U, Gubler DJ, Huang CYH, Burke DS, Mpoudi-Ngole E, et al. Seroprevalence and distribution of flaviviridae, togaviridae, and bunyaviridae arboviral infections

in rural Cameroonian adults. The American Journal of Tropical Medicine and Hygiene. 2006 jun;74(6):1078-83. Available from: https://ajtmh.org/doi/10.4269/ajtmh.2006.74.1078.

[16] J E Diallo M, Janusz K, Lewis R, Manengu C, Sall A, Staples. Rapid assessment of yellow fever viral activity in the Central African Republic; 2010.
